# Phenome-wide association study of ovarian cancer identifies common comorbidities and reveals shared genetics with complex diseases and biomarkers

**DOI:** 10.1101/2023.09.08.23295233

**Authors:** Anwar Mulugeta, Amanda L Lumsden, Iqbal Madakkatel, David Stacey, S. Hong Lee, Johanna Mäenpää, Martin K. Oehler, Elina Hyppönen

## Abstract

**Background:** Ovarian cancer (OC) is commonly diagnosed among older women who have comorbidities. This hypothesis-free phenome-wide association study (PheWAS) aimed to identify comorbidities associated with OC, as well as traits that share a genetic architecture with OC.

**Methods:** We used data from 181,203 white British female UK Biobank participants and analysed OC and OC subtype-specific genetic risk scores (OC-GRS) for an association with 889 diseases and 43 other traits. We conducted PheWAS and colocalisation analyses for individual variants to identify evidence for shared genetic architecture.

**Results:** The OC-GRS was associated with 10 diseases, and the clear cell OC-GRS was associated with five diseases at the FDR threshold (p =5.6×10^−4^). Mendelian randomisaiton analysis (MR) provided robust evidence for the association of OC with higher risk of “secondary malignant neoplasm of digestive systems” (OR 1.64, 95% CI 1.33, 2.02), “ascites” (1.48, 95% CI 1.17, 1.86), “chronic airway obstruction” (1.17, 95% CI 1.07, 1.29), and “abnormal findings on examination of the lung” (1.51, 95% CI 1.22, 1.87). Analyses of lung spirometry measures provided further support for compromised respiratory function. PheWAS on individual OC variants identified five genetic variants associated with other diseases, and seven variants associated with biomarkers (all, p ≤4.5×10^−8^). Colocalisation analysis identified rs4449583 as the shared causal variant for OC and seborrheic keratosis.

**Conclusions:** OC is associated with digestive and respiratory comorbidities. Several variants affecting OC risk were associated with other diseases and biomarkers, with this study identifying a novel genetic locus shared between OC and skin conditions.

## 1. Introduction

Ovarian cancer (OC) is a notorious gynaecological disorder, with only 44% of affected women surviving 5 years.^1^ Reducing mortality associated with OC remains a challenge as the poor understanding of the origin and biology of the disease limits opportunities to prevention, and the disease is typically detected at an advanced stage when treatment options are limited.^2^ Approximately 90% of OC are epithelial OC and arise from epithelial cells, predominantly of the fallopian tube and endometrium.^2–4^ Epithelial OC can be further classified into serous, endometroid, clear cell, and mucinous subtypes, based on their histological characteristics^3^ and indeed, the pathogenesis, disease progression, and risk factors are known to vary between OC subtypes.^3, 5^ Given that OC is commonly diagnosed in women aged 50 years and above,^6^ many patients present with at least one comorbidity, which can influence treatment decisions, impact prognosis and affect overall survival.^7, 8^ Hence, exploring the associations between OC and comorbidities, along with understanding potential shared genetic links between OC and other traits, may offer insights into the underlying etiology of the disease. Such insights could lead to facilitating earlier detection and better management of the disease.

Phenome-wide association study (PheWAS) is a powerful approach for testing the association of genetic variants or their risk score with a broad range of phenotypes and disease outcomes.^9^ This approach offers the advantage of being hypothesis-free, allowing it to explore numerous associations without constraints of the number of outcomes. Consequently, PheWAS has the potential to unveil novel genetic associations with diseases.^9^ In this study we use data on genetic OC risk variants and 889 disease outcomes from 181,203 female UK Biobank participants to identify comorbidities and clinical characteristics associated with the genetic risk of OC. We test for evidence of a direct association between OC and possible comorbidities using Mendelian randomisation (MR), an approach less susceptible to confounding and reverse causation,^10^ and extend these analyses to focus on the genetic risk reflecting distinct histological OC subtypes. We also use the PheWAS approach to explore each OC associated variant individually, which can provide evidence for shared genetic contributions, and provide insight into the underlying disease mechanisms.

## 2. Methods

### 2.1. Study population

UK Biobank is a prospective cohort study of half a million participants with rich phenotype and genotype data collected across 22 centres in England, Scotland, and Wales. The baseline assessment was conducted between 2006 and 2010 when the participants were aged between 37 to 73 years.^11^ A wide range of data was collected, including sociodemographic information, lifestyle factors, physical measures, blood and urine samples for biomarker and genetic profiling, with continuing updates through follow-up assessments and linkage with electronic health records and death registrations.^11^ For the primary analyses in this study, we restricted the sample to 181,203 female participants who were of white British ancestry and were not genetically related (Figure S1) and included male participants in sensitivity analyses (n =156,220). The study was conducted under UK Biobank application number 89630. UK Biobank obtained informed consent from all participants to collect and use linked data for future research use, and the UK Biobank study has ethical approval from the National Information Governance Board for Health and Social Care and Northwest Multicenter Research Ethics Committee (11/NW/0382).

### 2.2. Disease outcomes and biomarkers

We used International Classification of Diseases (ICD version 9 and 10) codes from hospital episode statistics and death registrations to identify disease outcomes (Supplementary methods). We mapped ICD-9/10 codes to phecodes, which are composite groupings reflecting clinically relevant disease outcomes.^12^ We defined individuals with the phecode-of-interest as cases, and those without the phecode-of-interest, or any other phecodes from the same disease category as controls (Supplementary methods).^12, 13^ We included all phecodes with at least 200 cases in our analyses,^14^ resulting in 889 phecodes from 18 disease categories (Table S1).

We also included 30 serum and four urine biomarkers, which are commonly used for the diagnosis and monitoring of chronic diseases, as outcome measures (Supplementary methods). Additional analyses were conducted against selected physiological characteristics (namely body mass index (BMI), body fat percentage (BFP), basal metabolic rate (BMR), blood pressures, and measures of lung function (spirometry) due to their known associations with comorbidity risks. The spirometry measures included forced expiratory volume in 1-second (FEV1), forced vital capacity (FVC), FEV1/FVC ratio, and peak expiratory flow (PEF).

### 2.3. Genetic variants and genetic risk score

We selected 35 genetic variants associated with OC from previous genome-wide association studies (GWAS), including the 12 novel variants identified in the most recent GWAS ^15^ and 23 variants identified in earlier GWASs (minor allele frequency ≥1.4%, Table S2, Figure S2).^16–23^ We constructed the genetic risk score (GRS) using 31 genetic variants after excluding one variant that had no proxy at r^2^ ≥ 0.8 (rs555025179), and three variants (rs12131772, rs4691139, and rs12938171) that were not replicated in the recent GWAS,^15^ which we extracted from the imputed genotype UK Biobank data ^24^ (Table S2). The GRS was constructed for each individual by summing the weighted number of OC risk-associated variants, with the weight taken from the recent OC GWAS.^15^ We also constructed risk scores for each OC subtype (Supplementary methods).

### 2.4. Statistical analyses

We conducted phenome-wide analyses using R-package PheWAS ^25^ which allowed us to fit logistic regression (889 disease outcomes) or linear regression (43 biomarkers/physiological characteristics) against the OC GRS with all models adjusted for age, assessment centre, type of genotyping array, birth location, and 40 principal components (Figure S2). We repeated the phenome-wide analyses for each OC subtype GRS to identify subtype-specific disease associations, and further, conducted the analyses in male participants to understand whether the OC – disease associations involve non-sex-specific mechanistic pathways.

The phenome-wide association signals that passed the FDR threshold (p <5.6 × 10^−4^ for the diseases and p <0.02 for the biomarkers and physiological measures, Supplementary methods) were taken forward to MR analysis to examine evidence for direct association with OC. We used two-sample MR approach where variant – OC association estimates were taken from the recent OC GWAS,^15^ and the variant-outcome association estimates were from the UK Biobank. MR analyses were conducted on signals associated with the overall OC GRS or with any subtype-based GRSs. We used inverse variance weighted (IVW) MR^26^ as the primary method and conducted sensitivity analyses using weighted median,^27^ weighted mode,^28^ MR-PRESSO,^29^ and MR-Egger,^30^ with each method working on different pleiotropic assumptions (Supplementary methods). We tested for horizontal pleiotropy using MR-Egger intercept, MR-PRESSO outlier and distortion tests, leave-one-out and leave-block-out methods (Table S3, Supplementary methods). MR analyses were repeated excluding the OC cases. To test the MR assumption that the genetic instrument should not be associated with potential confounding factors, we explored the association between the OC GRSs and age, education, Townsend deprivation index, BMI, physical activity, and alcohol consumption (Supplementary methods, Table S4).

We repeated the phenome-wide analyses for each OC genetic variant individually, again conducting analyses separately for men. We assessed the novelty of the PheWAS findings by checking the reported associations in the GWAS Catalog. We considered a locus-trait association as potentially novel if neither the lead variant nor any variant in linkage disequilibrium (LD) with the lead variant (with an r^2^ ≥0.1) had previously been reported to be associated with the same or related traits. For those potentially novel associations, we conducted a follow-up colocalisation analysis using the HyPrColoc (Hypothesis Prioritization in multi-trait Colocalisation) tool to assess whether there is evidence of shared causal genetic variants between pairs of traits.^31^ We considered a posterior probability of full colocalisation (PPFC) >0.64 as indicative of shared genetic etiology between these traits (Supplementary methods).^31^

Statistical analyses were performed using STATA SE version 17.1, R version 4.2.1 and PLINK2.^32^

## 3. Results

We included up to 181,203 female white British participants in the primary analyses, of whom 1,863 (1.3%) had been diagnosed with OC (Table 1). The prevalence of OC was higher among participants who were older, obese, never drinkers of alcohol, and those with poor general health status at the baseline (for all, prevalence ≥1.2%, and p ≤0.03). The OC risk GRS was normally distributed and associated with OC as expected, with stronger association in the highest compared to the lowest genetic risk group (F-statistics =23.7, p =4.4 × 10^−16^, Figure S3). No associations were found between the risk scores and potential confounders after correction for multiple testing (Table S5).

**Table 1.**
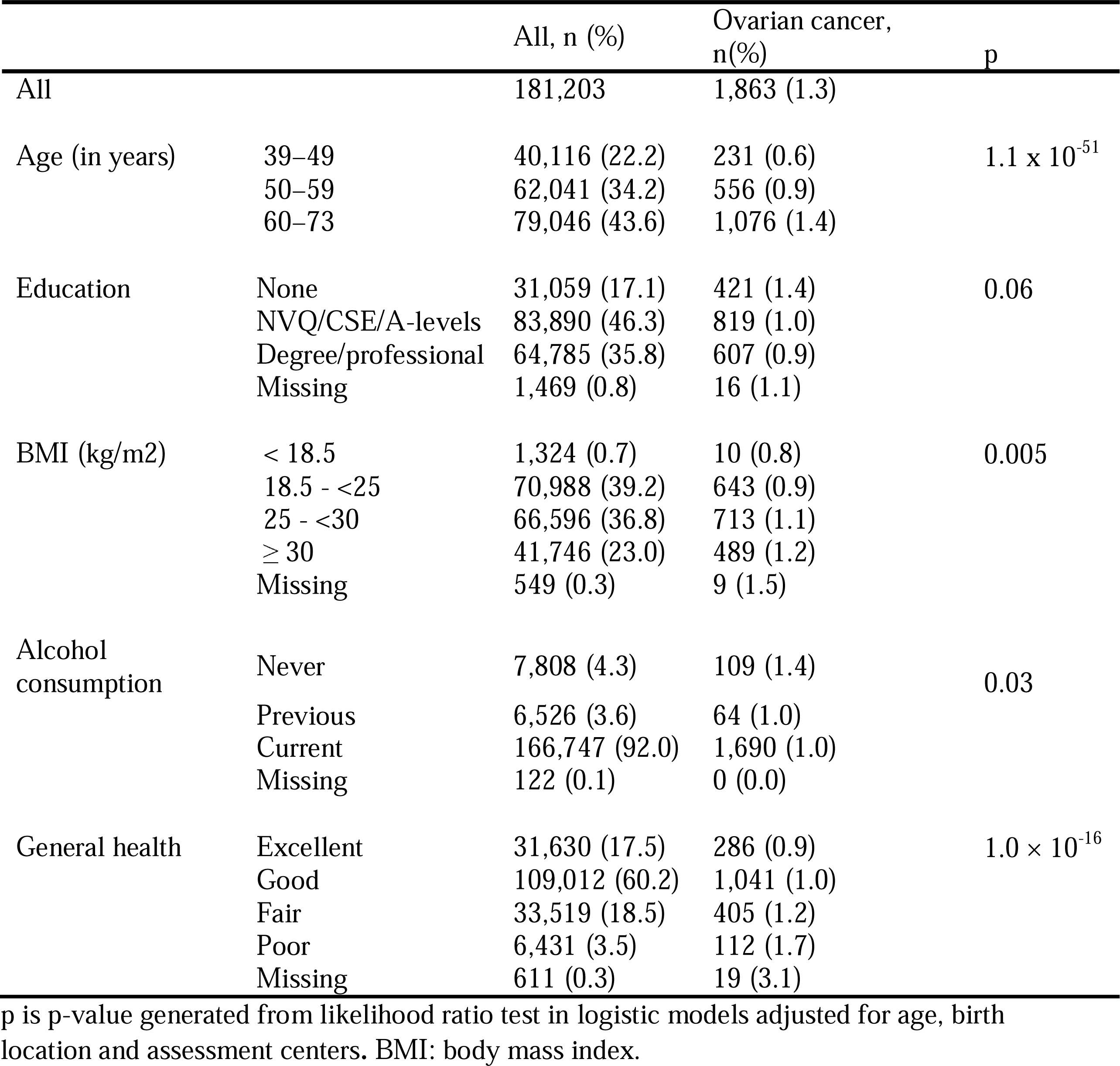
Prevalence of ovarian cancer by population characteristics.

### 3.1. Phenome-wide-based Mendelian randomisation

We investigated 889 disease outcomes from 18 disease categories in the phenome-wide analyses. We observed significant associations between the OC GRS and 10 diseases from four disease categories (neoplasms, digestive, respiratory and genitourinary diseases) after FDR correction (p ≤5.6 × 10^−4^). The top signals were for “cancer of the female genital organs”, “malignant neoplasm of ovary and other uterine adnexa”, and “malignant neoplasm of the ovary”, which confirms the ‘relevance assumption’ of the MR approach^10^ (Supplementary methods) for using the GRS as instrument for OC (for all, p ≤4.8 × 10^−4^, Figure 1, Panel A). The GRS was also associated with higher risk of other cancers and abnormal growths including “secondary malignant neoplasm of digestive systems”, “lipoma, including lipoma of skin and subcutaneous tissue” (p ≤3.9 × 10^−4^), and “ovarian cyst” (p =6.0 × 10^−5^). In addition, we identified associations with respiratory outcomes including “chronic airway obstruction” and “abnormal findings on examination of the lungs”, and with the digestive system outcome, “ascites” (p ≤3.2 × 10^−4^).

**Figure 1.**
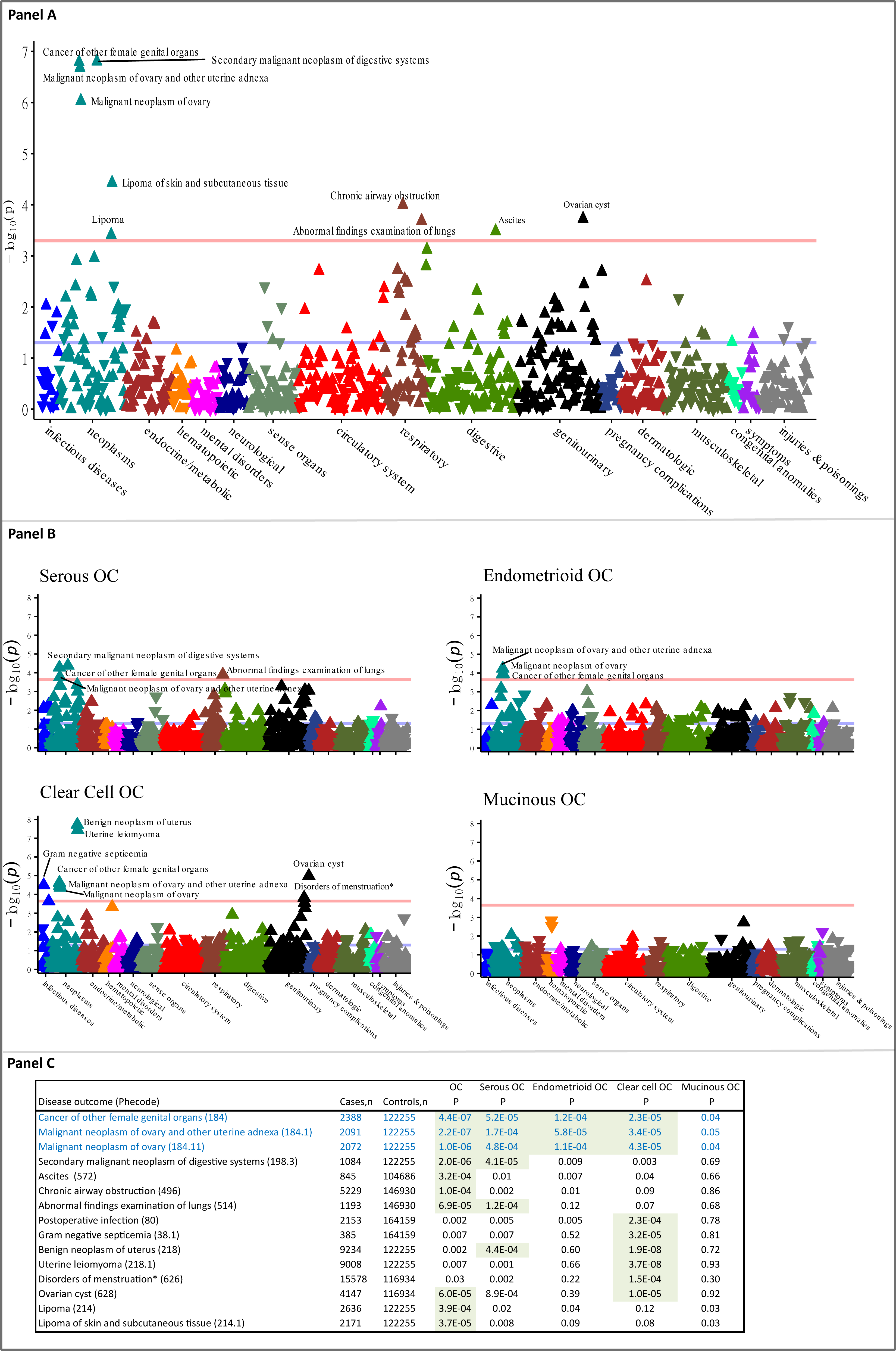
Manhattan plot showing the phenome-wide association of genetic risk scores of overall (Panel A) and subtypes (Panel B) of ovarian cancer with wide ranges of diseases. Panel C summaries the association of the top 15 signals identified at FDR threshold (p =0.0005) in Panel A or Panel B across the different genetic risk scores. The association for the first three diseases (in blue) is a validation of the genetic risk scores. Green highlights indicate association that passed the FDR threshold.

In analyses using GRSs for specific OC subtypes (Figure 1 Panel B and C), the clear cell OC GRS was associated with additional diseases and increased risks such as “benign neoplasm of uterus”, and “uterine leiomyoma”, “disorders of menstruation”, “gram negative septicemia”, and “postoperative infection” (for all, p ≤2.3 × 10^−4^). Overall, in addition to overall OC-related outcomes, 12 other disease outcomes were associated with at least one form of the OC GRS. Notably, none of these associations were detected when we repeated the phenome-wide analyses among men (Figure S4). Additionally, our phenome-wide analyses on 43 quantitative traits, which included serum and urine biomarkers and physiological measures, identified an association of OC GRS with 13 traits. Furthermore, we found an additional 11 traits associated with at least one subtype of OC GRS (Figure S5).

We conducted two-sample MR analyses on 12 disease outcomes identified in the PheWAS. For each outcome, evidence of causality was identified for at least one type of OC (Figure 2). We found robust evidence supporting the association between a higher genetic liability of OC and an increased risk of “secondary malignant neoplasm of digestive systems” (OR 1.64, 95% CI 1.33, 2.023), “ascites” (1.48, 95% CI 1.17, 1.86), “chronic airway obstruction” (1.17, 95% CI 1.07, 1.29), and “abnormal finding on the examination of the lungs” (1.51, 95% CI 1.22, 1.87). These associations were further supported by other MR methods (Figure S6). Additionally, we observed an association between a higher genetic liability of OC and an increased risk of “ovarian cyst” (1.23, 95% CI 1.04, 1.46), as well as “lipoma including lipoma of skin and subcutaneous tissues” (1.31, 95% CI 1.06, 1.61). Notably, the weighted median and weighted mode MR methods specifically supported the association with “lipoma of the skin and subcutaneous tissues” (Figure S6). There was some evidence supporting the association between OC and higher risk of “postoperative infection” and “gram-negative septicemia”, identified in the phenome-wide analyses on clear cell OC. However, these associations were observed only in the IVW and MR-PRESSO MR methods (Figure S6). In the MR analyses on OC subtypes (Figure 2), serous, endometrioid and clear cell OC were all associated with “secondary malignant neoplasm of digestive systems” and “ascites”. Serous and endometrioid subtypes showed evidence of effects on the respiratory outcomes. While the case numbers were lower for mucinous OC, this histotype was the only one to show evidence of an association with “lipoma of the skin and subcutaneous tissues” as evidenced by all MR methods. However, mucinous OC was not associated with any of the other outcomes (Figure S6). The clear cell subtype was associated with “ovarian cyst”, “benign uterine neoplasms”, “uterine leiomyoma”, and “disorders of menstruation”. Both serous and clear cell subtypes increased the risk of postoperative infection traits, with clear cell also increasing risk for “gram negative septicemia” (Figure 2 & Figure S6). We conducted analyses also excluding all known OC cases, and associations with “secondary malignant neoplasm of digestive systems” and “ascites” were attenuated, while estimates for all other outcomes were generally similar to the main findings (Figure S7). The MR-Egger intercept did not suggest evidence of pleiotropy for most associations, except for the association between serous OC and “postoperative infections” (p_intercept_ =0.01), which therefore should be cautiously interpreted. No evidence of pleiotropy was found using the MR-PRESSO outlier test and leave-one-out analysis (Figure S8). The estimates obtained from leave-block-out analyses were broadly consistent with the overall findings (Figure S9).

**Figure 2.**
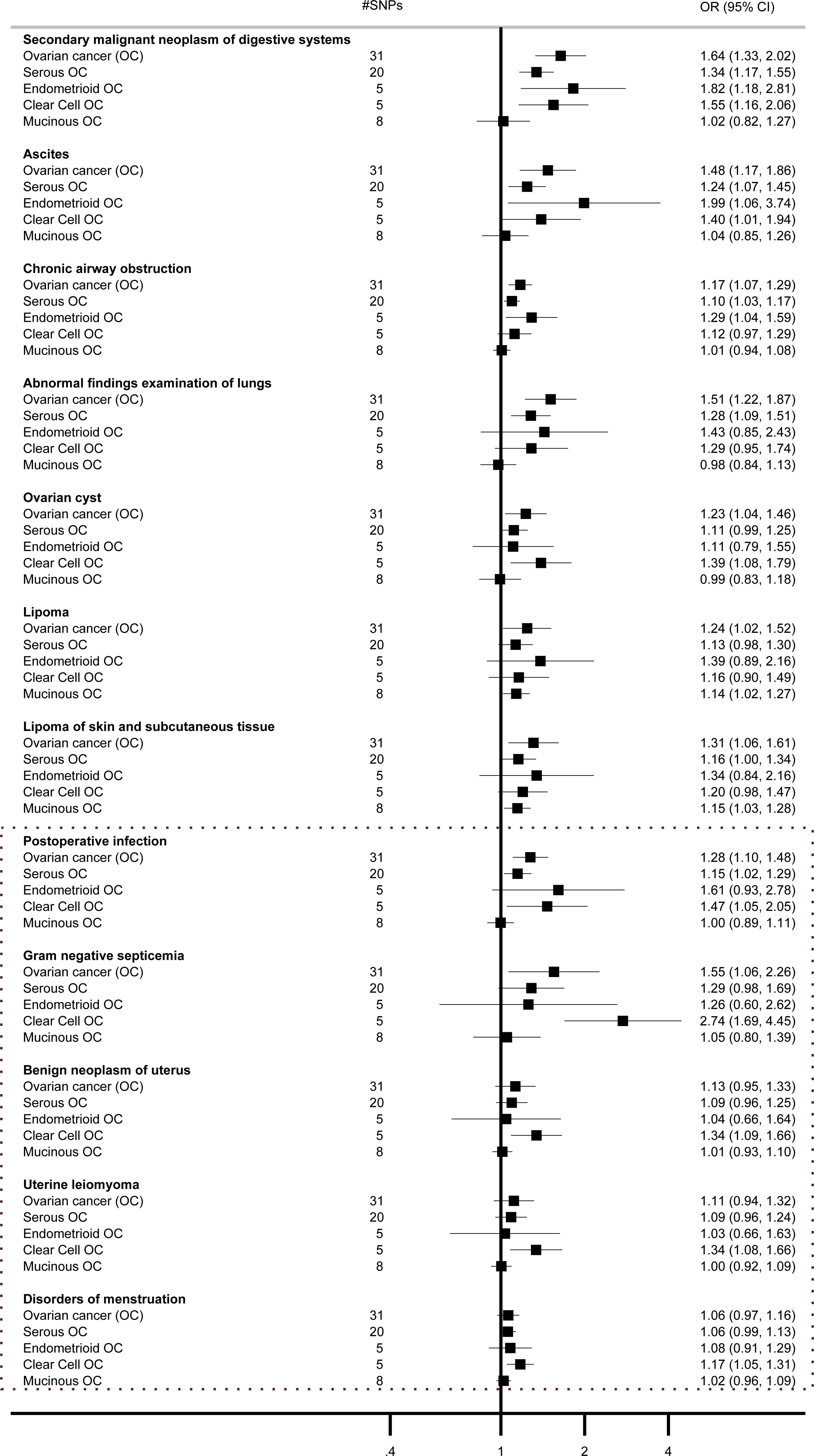
Mendelian randomisation causal estimates between ovarian cancer subtypes and selected diseases outcomes. Estimates were from Inverse variance weighted methods. #SNPs are number of genetic variants included in the analyses.

MR analyses on serum biomarkers and physiological measures supported the association of OC with lower FVC, endometrioid OC with lower oestradiol, clear cell OC with lower total cholesterol, low density lipoprotein (LDL) cholesterol, and apolipoprotein B (ApoB), and mucinous OC with higher gamma glutamyltransferase (GGT) and alanine aminotransferase (ALT) and lower creatinine levels (Figure S10). Analyses excluding all known OC cases, provided generally similar estimates to the main findings (Figure S11). For these identified associations, we did not find any evidence of pleiotropy based on the MR-Egger intercept tests (p_intercept_ ≥0.13) and leave-one-out analyses. MR analyses on men provided evidence for a genetic association between mucinous OC and ALT (Figure S10).

### 3.2. Shared genetic architecture

In the phenome-wide analyses assessing associations with individua OC risk-associated genetic variant, five variants were associated with at least one of 15 (11 distinct) disease outcomes (all, p ≤4.5 × 10^−8^, Figure 3, Table S6), and seven variants were associated with at least one of 22 biomarkers and physiological measures (Figure S12). Some of these associations were also observed in the analyses among men, with selected OC variants affecting conditions of the male genital organs (Figure 3, Table S6 and Figure S12, Supplementary Results). To ensure the novelty of our findings, we extensively checked the GWAS Catalog for any reports of associations with the identified (or related) diseases involving the lead or proxy variants (r^2^ ≥0.1). As a result, we uncovered potential novel associations between the OC risk-increasing allele from the *TERT* locus (rs10069690-T) and a lower risk of “seborrheic keratosis” (0.79, 95% CI 0.72, 0.87), in females, with similar a finding observed in males. To further validate the associations, we conducted colocalisation analysis at the *TERT* locus (LD block of chr5:982252 to chr5:2132442, human genomic build of hg19) and identified rs4449583 as the candidate causal variant explaining around 70% of 0.9239 PPFC for OC-“seborrheic keratosis” (Figure 4). This variant is also strongly associated with OC in the Phelan OC GWAS^15^ (p =7.8 ×10^−12^), and in LD with the index variant from this GWAS (rs7705526, r^2^ = 0.784) and with the lead variant in the primary discovery OC GWAS^22^ (rs100699690, r^2^ =0.432, Supplementary Methods). The association between the *GPX6* locus and “disorders of iron metabolism” and “celiac disease” are potentially novel, although colocalisation analysis did not suggest that OC share the same causal variants with these two diseases (Figure S13). On the contrary, we identified independent potential causal genetic variant for each “disorders of iron metabolism” (rs34409925, r^2^ = 0.207 with rs6456822) and “celiac disease” (rs3131101, r^2^ =0.049 with rs6456822) in further conditional analysis (Figure S13).

**Figure 3.**
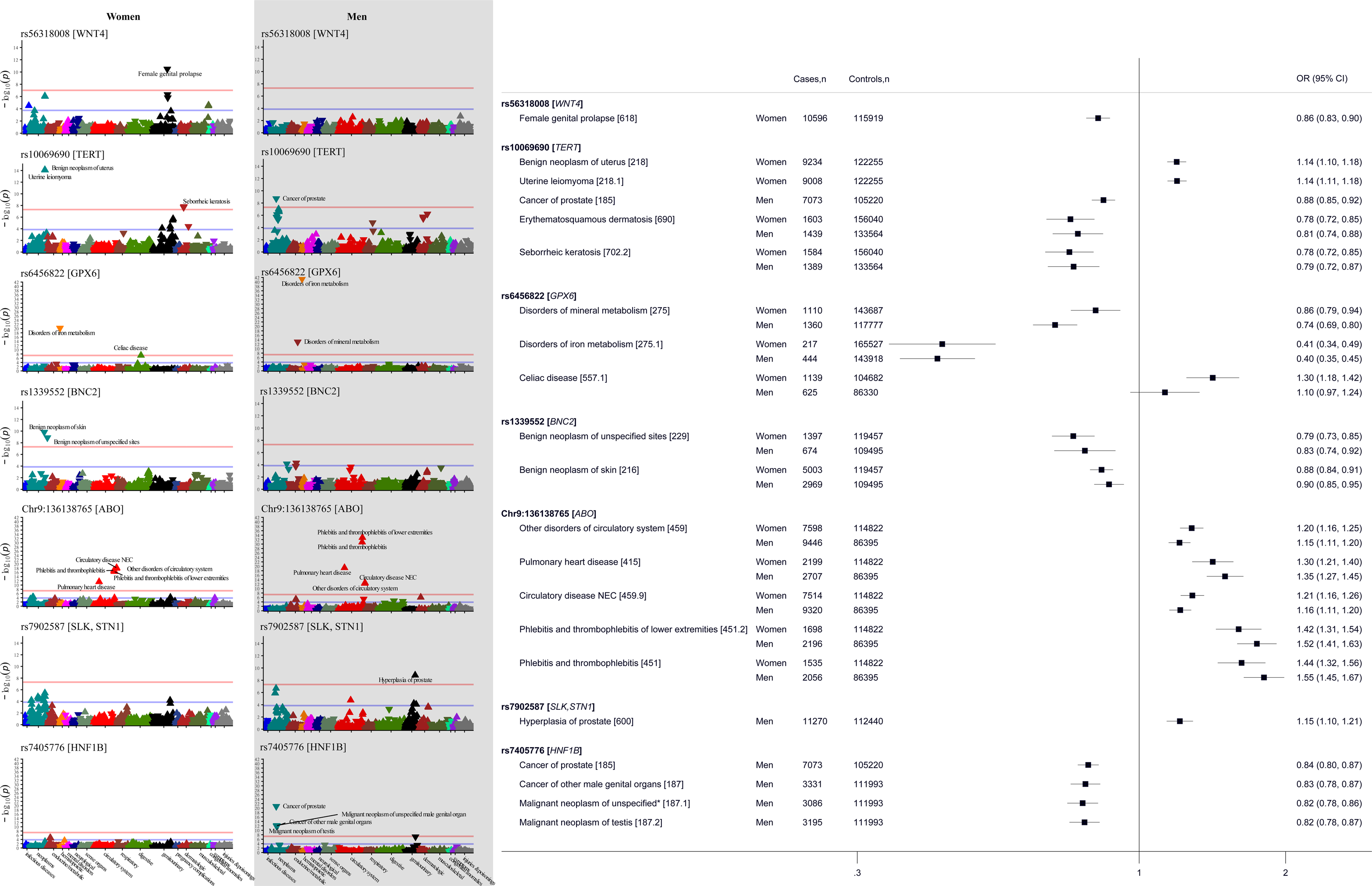
Phenome-wide association findings of ovarian cancer risk-increasing variants, with the Manhattan plot showing selected variants associations across all the diseases outcomes (with the y-axis showing the level of significance in −log_10_ scale), and the forest plot showing the selected variant – disease associations estimate among women and men.

**Figure 4.**
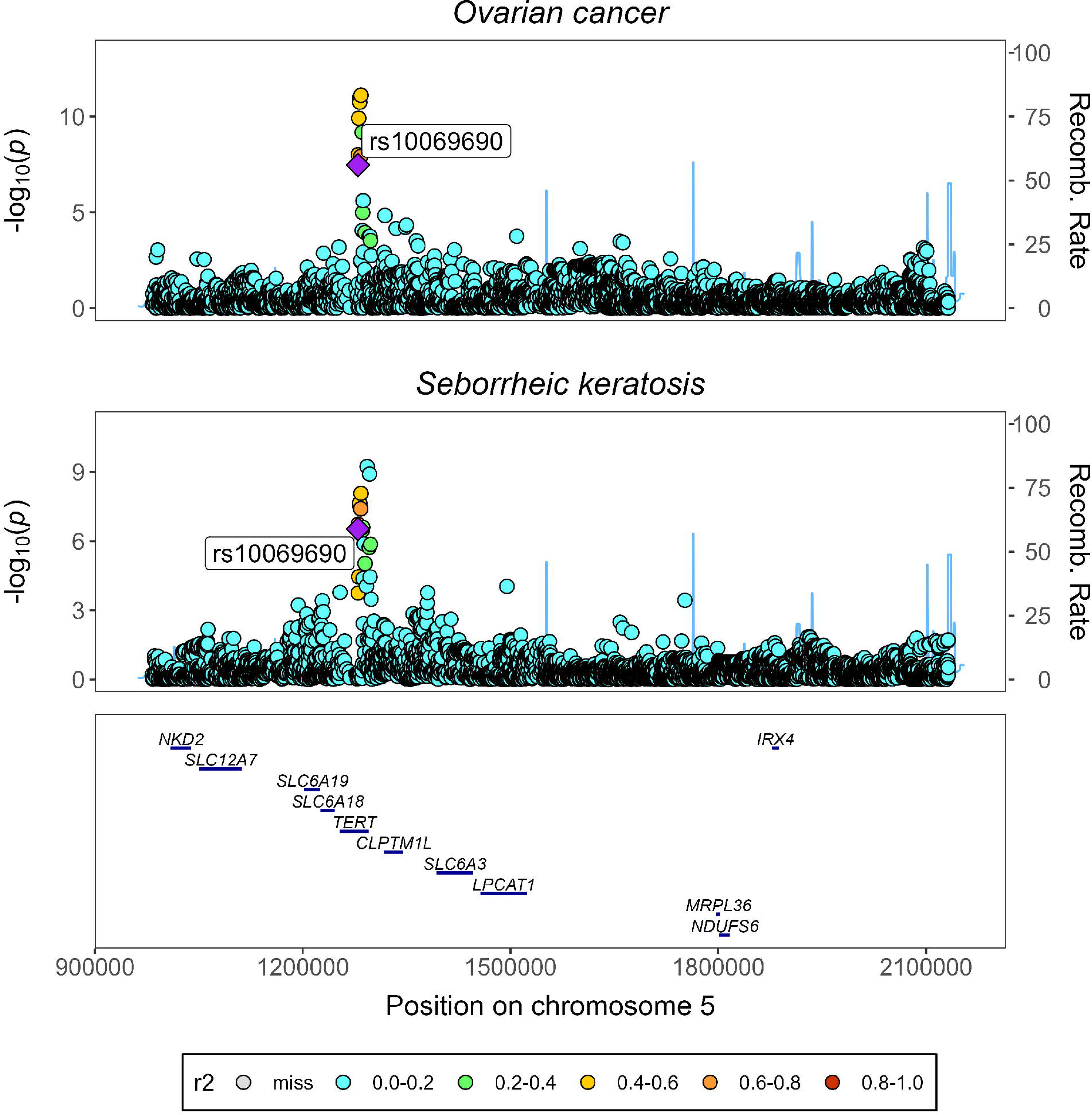
Colocalisation analysis of ovarian cancer with dermatological condition at the *TERT* locus. Panel A. Stacked regional association plots of ovarian cancer and seborrheic keratosis. Panel B. Portions of the Posterior probability for full colocalisation (PPFC) explained by each genetic variant at the *TERT* locus. The r^2^ colour legends reflect the r^2^ of each variant with respect to the lead variant.

## 4. Discussion

In this large-scale phenome-wide study, we aimed to explore comorbidity risks associated with OC, with additional analyses to identify subtype-specific associations based on genetic variants reflecting different OC histological types. We found evidence supporting a possible causal association between OC and 12 disease outcomes, including “secondary malignant neoplasm of the digestive system”, “ascites”, and respiratory disorders. We also identified some additional subtype-specific associations, including clear cell OC showing links with “gram-negative septicemia”, “uterine leiomyoma” and “disorders of menstruation”, and mucinous OC with “lipoma of the skin and subcutaneous tissue”. Analyses using 43 biomarkers also identified an association with seven biomarkers and these findings, along with those from disease analyses, may have important implications for improving the clinical management of patients with OC. Moreover, our exploration of each OC-risk increasing variant in the PheWAS revealed a set of traits that are genetically shared with OC. This analysis not only confirmed previous GWAS findings but also shed light on novel associations deserving further investigations.

OC is the leading cause of cancer-related ascites in females.^33^ Our MR-PheWAS study revealed genetic evidence linking ascites to OC across all subtypes except mucinous OC, which is consistent with the fact that over one-third of women with OC develop ascites during their lifetime.^33, 34^ Ascites is commonly considered a sign of advanced OC,^35^ and in line with this, the evidence for an association in our analyses was removed after excluding known OC cases. In patients with OC, the presence of ascites commonly indicates metastasis to peritoneal surfaces and reflects the extent of the disease.^36^ Our study further established a causal association between OC and “secondary malignant neoplasms of the digestive system”, indicating intra-abdominal dissemination of the tumour. Again, this association was sensitive to the inclusion of known OC cases in our analyses, and is likely to explained by a metastasis of the OC affecting the gastrointestinal tract or, potentially, a new synchronous gastrointestinal cancer.

OC has also been linked to respiratory complications.^37, 38^ Our study provides additional supporting evidence for this association, specifically linking OC to an increased risk of “chronic airway obstruction” and “abnormal lung examinations". The findings were further supported by spirometry measures, which showed a significant association between a higher genetic liability to OC and lower FVC. While in women with OC lowering of lung function may follow from malignant pleural effusion and tumour metastasis to the pleura^37^ or complications related to surgical procedures,^38^ in our analysis the association with lung disease remained when OC cases were excluded, and the spirometric association was seen both in women and in men, which may indicate a more profound biological link.

OC as reflected by our genetic study arises from the epithelium of the ovaries, endometrium and fallopian tubes, and while most of the lung also consists of epithelium, a direct link has not been well established. Interestingly, colocalisation analyses in our study also suggested that disorder of skin condition (seborrheic keratosis) shared a genetic locus (*TERT*, encoding the enzyme, telomerase) with OC, highlighting potential shared pathomechanisms involving the epithelium, and implicating telomerase function in the opposing risks of OC and the skin disorder.^39, 40^

Debulking cytoreductive surgery is the standard procedure for advanced OC. It involves the surgical removal of female reproductive organs such as the ovaries, fallopian tubes, uterus, as well as omentum and organs of the digestive system, such as the bowel, depending on the stage of OC.^41^ The invasive nature of this intervention,^41^ combined with compromised immune system function resulting from the disease,^42^ may explain the observed association between OC and “postoperative infection” and ”gram-negative septicemia”. It is important to note that these associations were predominantly observed in clear cell OC, which has also been associated with “benign neoplasms of the uterus”, “uterine leiomyoma”, and “disorders of menstruation”. The presence of these conditions, which primarily involve the uterine tissue, could reflect the well-established association between endometriosis and clear cell OC.^43, 44^ Our study also identified OC subtype specific associations between mucinous OC and “lipoma of the skin and subcutaneous tissue”. Regarding the biomarkers, endometroid OC was associated with lower serum oestradiol level, clear cell OC was associated with lower serum total cholesterol, LDL cholesterol, and ApoB, while mucinous OC was associated with higher serum GGT and ALT as well as lower creatinine levels. These findings suggests that comorbidities and selected biomarkers may play a role in identifying and characterising different subtypes of OC.

Beyond the causal link between OC and disease outcomes and biomarkers, genetic variants contributing to a higher risk of OC may also be associated with other diseases and biomarkers. The *ABO* locus is a well-known pleiotropic locus with established associations with cardiovascular and cerebrovascular traits.^45–50^ Our study confirmed these existing findings, as we also observed associations between the *ABO* variant and conditions such as “pulmonary heart disease”, “thrombophlebitis”, and “other disorders of circulatory system”. Additionally, we confirmed the association between *ABO* with many cardiometabolic biomarkers, including associations with markers of glucose and liver metabolism, confirming the highly pleiotropic nature of this variant.^51^

Our study identified a significant association between the *GPX6* variant (rs6456822) and a lower risk of disorders related to iron metabolism, which is a critical component of overall health.^52^ However, while the lead variant rs6456822 showed this association, our colocalisation analysis identified an independent potential causal variant, rs34409925, for “disorders of iron metabolism”. We also found another potential causal variant at the *GPX6* locus, rs3131101, for “celiac disease”. The OC lead variant rs6456822 was also associated with total protein levels in women, as well as with SHBG levels in men. Our study also identified a potential novel association between the *TERT* locus (rs10069690) and “seborrheic keratosis”. The follow up colocalisation analysis strengthened the shared genetic structures between this skin condition and OC, further suggesting that rs444583 may be a causal variant. Importantly, we confirmed previous associations of the *TERT* variant with uterine leiomyoma^44, 53^ and total protein levels in women^49, 51^, as well as with prostate cancer in men.^54^ Our findings highlight the value of using PheWAS for validating previous GWAS findings and identifying potential novel variants for multiple traits including diseases and biomarkers that never been tested before.^55^

Our study has a number of strengths. Firstly, it is the largest (n =181,203) and most comprehensive study to date that examines the health risks associated with a higher genetic susceptibility to OC. Secondly, by including OC subtypes in our analysis, our study can identify subtype-specific comorbidities, supporting more effective treatment and management. Lastly, our study extended the phenome-wide analyses to each OC-risk-increasing variant. This approach enables us to confirm previously identified variant-disease associations and also identify potential novel associations, thus providing a more comprehensive understanding of the shared genetic links between OC and other diseases. Further incorporation of colocalisation analysis is another strength of this study. It provided evidence of a shared genetic locus (*TERT*) between OC and “seborrheic keratosis”, and identified a potential novel causal variant, rs4449583, for this skin condition.

Our study also has some limitations. To ensure the validity of MR, the genetic instrument used must be associated with the risk factor (in our case, OC), not associated with the confounders of the risk factor-outcome association, and not influence the outcome except through its association with the risk factor. While we used a genetic instrument that associates with OC and conducted our analysis on a white-European population while adjusting for confounding factors, we cannot rule out the absence of horizontal pleiotropic effects, which will affect interpretation and can bias the MR estimates.^56^ To address this concern, we repeated our analysis in men to further verify whether the association we observed is genuinely related to OC or a reflection of genetic variants indicating shared mechanistic pathways between both men and women. Another potential limitation of our study is that the UK Biobank participants are relatively healthy compared to the general population,^57^ introducing to collider bias due to the "healthy volunteer" selection bias.^58^ However, the modest collider bias in the context of MR is less likely to affect MR results significantly when the selection into the study was not strongly influenced by the risk factor.^58^ Furthermore, the presence of biologically plausible causal associations in our study provides support for the validity of our results despite this limitation. It could be noted as a limitation of our study that our investigation into comorbidities associated with OC was primarily conducted in a population predominantly not diagnosed with OC. However, sensitivity analyses excluding all known OC cases confirmed that the associations with OC complications relating to ascites and gastrointestinal metastases are related to the disease *per se*, while the other observed disease and trait associations may reflect earlier stages of OC development or other underlying biological differences.

In conclusion, our study identified robust associations between OC and comorbidities related to the digestive and respiratory systems. Furthermore, we observed specific robust associations for different subtypes of OC, such as “gram-negative septicemia”, ‘uterine leiomyoma” and “disorders of menstruation” for clear cell OC, and lipoma for mucinous OC. Interestingly, our study points to a shared genetic architecture between OC and a skin condition, leading to a discovery of a novel variant which merits further investigation.

## Funding

This study was supported by Medical Research Future Fund of Australia (MRFF), grant number 2007431.

## Competing interests

The authors declare that they have no competing interests.

## Acknowledgements

The authors are very grateful to the UK Biobank and UK Biobank Participants.

## Authors’ contributions

AM conceived the study with EH, conducted the analysis, and wrote the first draft. E.H. supervised the study and advised on analyses. DS conducted the colocalisation analysis using summary results generated by AM. All authors interpreted the results, revised the manuscript, and approved the final manuscript.

## Ethical approval

The study was conducted under UK Biobank application number 89630. UK Biobank study has ethical approval from the National Information Governance Board for Health and Social Care and Northwest Multicenter Research Ethics Committee (11/NW/0382).

## Consent to participate

UK Biobank obtained informed consent from all participants to collect and use linked data for future research use.

## Data availability

This research has been conducted using the UK Biobank Resource under Application Number 89630. The data and analytical code that support the findings of this study are available on application to the UK Biobank (https://www.ukbiobank.ac.uk/enable-your-research/apply-for-access).

---

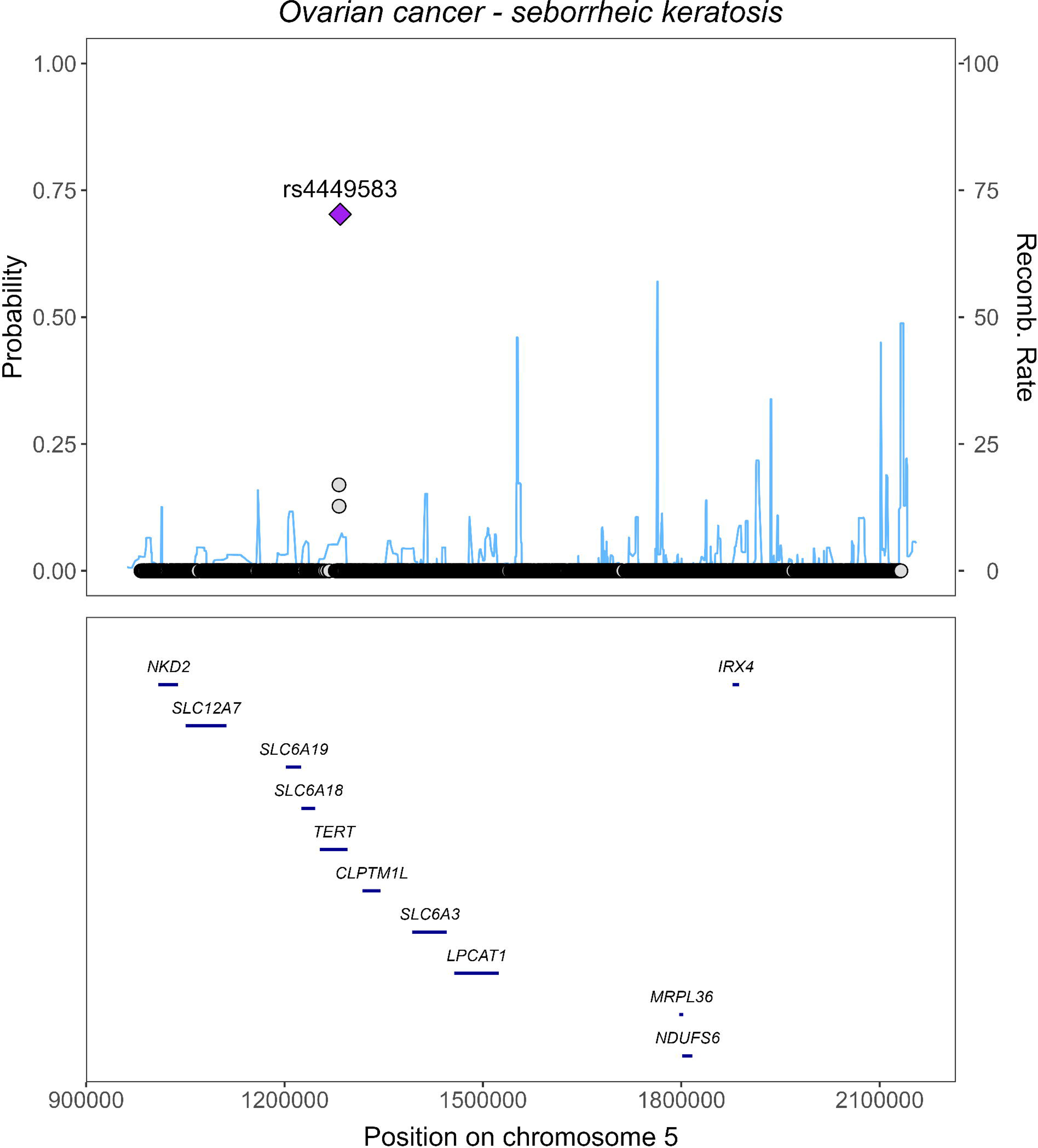

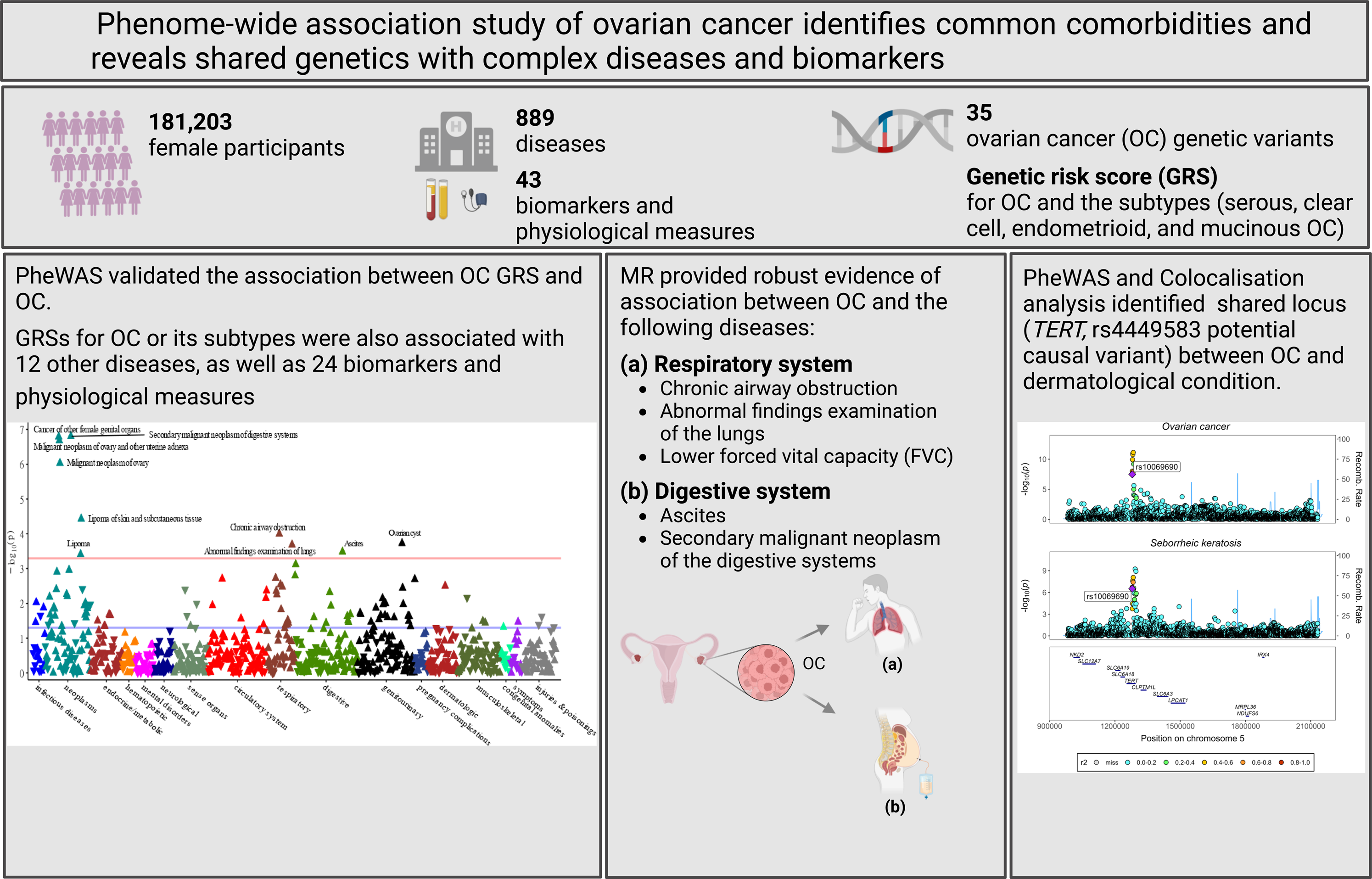

## Notes

### Competing Interest Statement

The authors have declared no competing interest.

### Author Declarations

UK Biobank obtained informed consent from all participants to collect and use linked data for future research use, and the UK Biobank study has ethical approval from the National Information Governance Board for Health and Social Care and Northwest Multicenter Research Ethics Committee (11/NW/0382).

