## Supplementary for "Phenome-wide association study of ovarian cancer identifies common comorbidities and reveals shared genetics with complex diseases and biomarkers"

**Short title**: MR-PheWAS of ovarian cancer

**Funding**: This research was funded by Medical Research Future Fund of Australia (MRF2007431).

***Corresponding author**:
Dr Anwar Mulugeta
Australian Centre for Precision Health, c/o SAHMRI, University of South Australia, Adelaide, Australia, GPO Box 2471, Adelaide SA 5001.


### Supplementary methods

#### Disease outcomes

The primary outcomes of the study included disease endpoints defined using ICD-9/10 (International Classification of Diseases version 9 and 10) codes recorded from the hospital episode statistics (inpatient data) and death registration. We used previously validated case-control mapping file to define the phecodes,[1, 2] which are disease endpoints with carefully curated cases and controls resembling those used in clinical and genomic studies.[3] Cases included individuals with phecode-of-interest. For instances, individuals with diagnosis for pulmonary embolism and infarction, primary pulmonary hypertension, and chronic pulmonary hypertension contributed to the pulmonary heart disease case definition. Controls included individuals without all cases within the disease category, as described before.[2] For example, for pulmonary heart disease, participants with any circulatory diseases diagnose would be excluded from the controls. We have defined 889 phecodes after excluding those with case number below 200 which is a recommended case number to achieve adequate power in genetic studies using common variant.[4] Excluding phecodes with case number below 200 may also minimise the multiple test burden. This case threshold was chosen as it has been shown to provide ≥80% power in genetic analysis using common variants (minor allele frequency >0.01, genetic penetrance ≥0.15, logOR =0.1, α =0.00025).[4]

To examine the distribution of ovarian cancer cases across different population characteristics and to further validate the genetic instrument, we utilized OC cases identified through information obtained from hospital episode statistics (inpatient data) and the cancer registry. Cases of OC were identified using ICD-10 (ICD-9) codes C56 (183.0) for malignant ovarian cancer, C57.0 (183.2) for malignant fallopian tube cancer, and C48.2 (158.9) for malignant peritoneal cancer. The control group consisted of individuals who did not have a history of cancer, including self-reported cancer information at baseline, with the exception of cases of non-melanoma skin cancer (C44.0-C44.9), benign tumors (D10-D36), and in-situ cancers (D00-D09). For the purpose of further subtyping OC, we relied on tumor behavior and histology information available from the cancer registry. Serous OC was defined by specific histological characteristics, including serous cystadenocarcinoma (code: 8441), papillary serous cystadenocarcinoma (8460), and serous surface papillary carcinoma (8461). Endometrioid OC was defined based on histological characteristics such as endometrioid carcinoma (8380), adenosquamous carcinoma (8560), and adenocarcinoma with squamous metaplasia (8570). Clear cell OC was defined by the presence of clear cell adenocarcinoma (8310). Lastly, mucinous OC was defined by histological characteristics such as mucinous cystadenocarcinoma (8470), papillary mucinous cystadenocarcinoma (8471), mucinous adenocarcinoma (8480), and mucin-producing adenocarcinoma (8481).

#### Biomarkers and physiological measures

As secondary outcomes, our study included serum and urine biomarkers, and physiological measures. For classification of biomarkers in different groups, we used the terminology used by the UK Biobank biomarkers Enhancement group.[5] Accordingly, our outcomes included cancer biomarkers (oestradiol, testosterone, sex hormone binding globulin (SHBG) and insulin-like growth factor-1 (IGF-1)), cardiovascular biomarkers (total cholesterol, low density lipoprotein (LDL) cholesterol, high density lipoprotein (HDL) cholesterol, triglycerides, apolipoprotein A1 (ApoA1), apolipoprotein B (ApoB), lipoprotein A (Lp(A)), and C-reactive protein (CRP)), diabetes biomarkers (glucose and glycated haemoglobin (HbA1C)), renal biomarkers (serum creatinine, creatinine in urine, cystatin C, phosphate, total protein, urate, urea, potassium in urine, sodium in urine, microalbumin in urine), liver biomarkers (alanine aminotransferase (ALT), albumin, aspartate aminotransferase (AST), gamma glutamyltransferase (GGT), direct bilirubin and total bilirubin), and bone and joint biomarkers (alkaline phosphatase, calcium and vitamin D). Serum and urine samples were collected almost for all participants at the baseline and were further assayed centrally by the UK Biobank Biomarker enhancement groups. Most of the serum biomarkers assayed using Beckman Coulter AU5800 analytical platform, while Cystatin-C used Siemens Advia 1800, IGF-1 and vitamin D used Diasorin Liaison XL, and Oestradiol, testosterone and SHBG used Beckman Coulter DXI 800.[5] Depending on the serum biomarkers different analytical methodologies including colorimetric, enzymatic rate, immuno-turbidimetric, enzyme immune-inhibition, enzyme selective protection, and chemiluminescent immunoassay were utilised for the respective assays, and details can be found elsewhere.[5] For urine biomarkers, all were assayed using Beckman Coulter AU5400 clinical chemistry analyser Beckman, with enzymatic analytical method used for creatinine, immune-turbidimetric for microalbumin and ion selective electrode method for potassium and sodium.[6] For serum biomarkers, we excluded data from aliquot three due to poor quality.[5] Measures for oestradiol, rheumatoid marker, and microalbumin in urine have >70% of values below the lower limit of detection (LLOD), which we replaced with LLOD/2 for all participants who undertake these assays. we used the values of 36.5 pmol/L for oestradiol, 5 IU/ml for the rheumatoid marker, and 3.35 mg/L for microalbumin. We applied log-transformation to biomarkers and physiological measures to mitigate the skewness of the distribution, and further scaled to a mean of zero and a standard deviation (SD) of one to allow us to present the results using the same scale for all measures.

We also included the baseline physiological measures such as body mass index (BMI, defined as weight over height (in m) squares), body fat percentage (BFP), basal metabolic rate (BMR), diastolic and systolic blood pressures (DBP and SBP)), and lung spirometry measure. The spirometry was measured using a Vitalograph Pneumotrac 6800 for from all participants at the baseline.

We included forced expiratory volume in 1- second (FEV1) and forced vital capacity (FVC), the ratio of these values (FEV1/FVC), and peak expiratory volume (PEF) for the current study.

#### Covariates

In addition to the basic covariates such as age and sex, we included different socioeconomic and lifestyle factors for characterising the study population and validation of genetic instrument. List of these covariates are described in the Table S4.

#### Genetic data

We extracted the genetic variants from imputed genotype UK Biobank data, with the genetic data were processed and checked for quality by the UK Biobank central genetic team.[7] UK Biobank used Axiom® platform, which contains over 800,000 genetic markers, for genotyping half a million UK Biobank individuals. In practice, they used two very similar genotyping panels which the UK BiLEVE array was used approximately 50,000 samples and UK Biobank Axiom Array for approximately 450,000 samples. The genetic marker information was maximised to over 96 million through imputation of the genotype missing variants using the Haplotype Reference Consortium as well as combined UK10K and 1000 Genomes Phase 3 reference panels.[7]

For identifying the OC associated variants, we search GWAS Catalog[8] for genome-wide association studies (GWAS) on OC and identified 10 GWAS conducted mainly in White European population (Figure S1 & Table S1). From these studies, 35 genetic variants with association to overall or subtypes of OC were identified at genome-wide significant threshold.[9-17] Of which, 12 novel variants were identified in the recent GWAS[18] while the other 23 variants were first identified in the earlier GWASs.[9-16] Of the variants, 26 were genome-wide significant in the recent GWAS,[17] and five of the earlier identified variants were replicated at nominal significance ($p < 0.05$) for a directionally consistent association with overall or a subtype of OC. Given the recent GWAS has maximum cases number compared with the earlier GWASs and did not include UK Biobank, we based our analysis using the weight taken from this GWAS.[17] In addition to the 12 novel loci identified in the recent GWAS, we included previously identified variants with an association with OC or its subtypes at least at nominal significant threshold ($p<0.05$) in the recent GWAS. Accordingly, three variants (rs12131772 [*AK5]*, rs4691139 [*TRIM60*, *TRIM61*], and rs12938171[*BRIP1*]) that were not replicated in the latest GWAS at nominal threshold were excluded (Table S1). We also excluded rs555025179 (or proxy at $r^2 \geq 0.8$, *MAST4*) which was not found in UK Biobank (Table S1). Of the included variants, 20 were associated with serous ovarian carcinoma, five with endometrioid carcinoma, five with clear cell carcinoma and eight with mucinous carcinoma in the recent GWAS 17 (Table S2). We coded each OC risk increasing alleles as 0,1 or 2 using additive mode.

#### Genetic risk score

We constructed the weighted genetic risk score (GRS) for overall and subtypes of OC using the equation below.

$$GRS = \frac{(\beta_1 \text{ x variant}_1 + \beta_2 \text{ x variant}_2 + \ldots \beta_n \text{ x variant}_n) * \text{total available number of variants}}{\text{Sum of } \beta \text{ coefficients}}$$

*Equation 1*

Where, β is the log-odds of the variant-OC (or subtypes) association from the recent GWAS, n is the number of genetic variants included in the risk score. The score was scaled by the ratio of available number of genetic variants and sum of beta coefficients. Weight for overall and subtype of OC was taken from recent GWAS from the analyses specific to each OC[17].

#### Statistical analysis

Our analyses were divided into two main sections. The first involved phenome-wide screening for diseases and biomarkers potentially associated with OC and its subtype GRSs, followed by MR to evaluate the robustness of OC-disease/trait association for the identified signals. The second involved conducting phenome-wide analyses for each genetic variant, identifying potential novel associations using the National Human Genome Research Institute (NHGRI)-European Bioinformatics Institute (EBI) Catalog (here after referred as GWAS Catalog),[8] and confirming the shared genetic architecture using colocalisation analysis (Figure S2). The statistical analyses were performed using STATA SE version 17.1 software, R version 4.2.1 and PLINK2.[19], with the analyses further utilised R-packages including PheWAS,[20] TwoSampleMR,[21] MR-PRESSO,[22] and HyPrColoc.[23]

##### Multiple test correction for the phenome-wide analyses

We conducted phenome-wide analyses on 889 phecodes, and 43 biomarkers and physiological measures to identify potential disease, clinical and physiological signals associated with OC Given the number of tests performed and interrelatedness of the outcomes (e.g., overall diabetes, type 1 diabetes, type 2 diabetes), we employed FDR correction for identifying the top signals. When we first screen for OC GRS against the phecodes, we identified 10 disease signals that passed the multiple test correction FDR threshold ($P < 5.6 \times 10^{-4}$, 10 identified diseases *0.05/889).[24] Additional diseases were identified from OC subtypes GRS phenome-wide scan using the same FDR threshold ($p < 5.6 \times 10^{-4}$). Similarly, for the biomarkers and physiological measures, we first screened for OC GRS at FDR threshold (p <0.02, 13 signals * 0.05/43), and add additional signals from the OC subtype GRS analyses at the same threshold.

**Mendelian randomisation (MR) methods and the assumptions**

Mendelian randomisation is a genetic instrument-based causal approach that applies three basic assumptions for providing the evidence of causality. Firstly, the genetic instrument should associate with the exposures (here, with OC, 'relevance assumption'). Secondly, there should not be association between the genetic instrument and the confounders of the exposure-outcome associations ('independence assumption'), and thirdly, the genetic instrument-outcome association should only be through the effect on the exposure ('exclusion restriction assumption').[25] We used multiple MR methods that works on different pleiotropic assumptions, and consistency across the methods suggesting the robustness of the findings. Random IVW model gives unbiased estimates if there is no or directionally balanced horizontal pleiotropy,[26] while weighted median remains valid estimator even up to 50% of the variants are pleiotropic.[27] Weighted mode gives valid causal estimates even if most of the instruments are pleiotropic as long as the most common (mode) horizontal pleiotropic effect is zero.[28] MR-Egger is most pleiotropy relaxing but more power demanding MR estimator. MR-Egger gives a valid estimate even if all the instruments are pleiotropic as long as the InSIDE (Instrument Strength Independent of Direct effects) assumption holds[29]. In principle, the MR-Egger intercept is unconstrained to zero, and the extent of deviation from zero, as indicated by the MR-Egger intercept test, suggest the presence of horizontal pleiotropy[29]. MR-PRESSO incorporates tests that allows to identify the potential pleiotropic variants if there is any (MR-PRESSO outlier test), and further examines whether the causal estimates varied before and after excluding the pleiotropic variants (MR-PRESSO Distortion test). We used leave-one-out and leave-block out analyses to further check the effect of pleiotropy on causal estimates. The leave-one-out analysis generates causal estimates by excluding each of the genetic instruments one at a time, while the leave-block-out analysis involves composition of functional blocks based on the association of the genetic variants (or proxy variants, $r2 \geq 0.8$) with other traits at genome-wide significance as identified from the GWAS Catalog,8 and then excluding each set of genetic variants of a given functional block one at a time to generate the MR estimates (Supplementary methods). For the leave-block-out analyses, we divided the variants according to eight functional blocks: cancers (not including OC), hormonal and gynaecological, anthropometric, cardiometabolic, biomarkers, blood count, respiratory, and miscellaneous (Table S3)

**Colocalisation analysis for shared genetic architecture**

We conducted colocalisation analysis for the OC locus-trait associations, which we considered a potential novel type after confirming the absence of this association for the lead or proxy variant with the given trait in the GWAS Catalog.[8] In our study, we used HyPrColoc, a Bayesian algorithm that can test multiple traits simultaneously to identify traits that colocalize at specific genetic loci.[23] This algorithm assumes that the genetic association across traits is collected from non-overlapping populations with similar population ancestry and LD structure.[23] In our phenome-wide association analysis, we identified the *GPX6* and *TERT* loci as potential novel loci for "disorders of iron metabolism" and "celiac diseases", and "seborrheic keratosis", respectively. Therefore, we selected these two loci along with these associated diseases for colocalisation analysis to provide evidence for shared genetic architecture with OC. For each locus, we confined the genetic variants window to the LD block, which was defined as Chr6:28017819 – 28917608 for the G6PD locus and Chr5:98252 – 2132442 for the TERT locus. The LD block was established using the 1000 Genome data and the hg19 human genome reference build.[30] We obtained the summary results for the association of the genetic variants within the target LD block and OC from the Phelan OC GWAS (please note the lead variant from *TERT* locus in this GWAS is rs7705526,[17] unlike the lead variant we used in our study, rs100699690, which is reported in the primary discovery GWAS.[15]). We generated the respective associations with the selected traits from the UK Biobank, with the selected disease outcomes based on the Phecode classification algorithm using the PheWAS package (see 'Disease outcomes' section above). We collected the results from logistic regression adjusting for age, centre, genotyping array, and 40 principal components, applying the PLINK analytical software. The probability of full colocalisation, known as the posterior probability for full colocalisation (PPFC), was calculated using two factors: the regional association probability, which is the probability that all the traits have a shared association with at least one genetic variant in the region, and alignment probabilities, which represent the likelihood of alignment at a single causal variant among the shared associations. The equation for calculating PPFC is shown below[23]. We employed a PPFC threshold of >0.64 to determine the colocalisation of traits at the locus.

$$PPFC = P_R * P_A,$$

Where $P_R$ is regional association probability, and $P_A$ is alignment probability.

**Supplementary Results**

The phenome-wide analysis using the individuals OC-risk variants identified the association of five variants with at least one of the 15 (11 distinct) disease outcomes (all, p ≤4.5 × 10-8, Figure 3, Table S6), and seven variants were associated with at least one of 22 biomarkers and physiological measures (Figure S12). By effect size, the strongest association estimates were observed for the rs6456822-T allele at the GPX6 locus, where each additional risk allele was associated with lower odds of "disorders of iron metabolism" both among women and men (OR 0.41, 95% CI, 0.34, 0.49 and 0.40, 95% CI 0.35, 0.45, respectively). A well-known pleiotropic variant in the ABO locus (Chr9:136138765) was associated with higher risk of circulatory diseases including "pulmonary heart diseases" (1.30, 95% CI 1.21, 1.40), "phlebitis and thrombophlebitis of lower extremities" (1.44, 95% CI 1.32, 1.56), and "circulatory diseases NEC" (1.21, 95% CI 1.16, 1.26) in women, with consistent genome-wide significant findings among men. We found associations between the OC risk-associated variant of rs10069690 (TERT) and "benign neoplasm of the uterus" (1.14, 95% CI, 1.10, 1.18), and also with lower risk of "prostate cancer" (0.88, 95% CI 0.85, 0.92) among men. The rs56318008-T allele (OC risk-increasing allele) from the WNT4 locus was associated with lower risk of "female genital prolapse" (0.86, 95% CI 0.83, 0.90) and the rs1339552-C allele at the BNC2 locus was associated with lower risk of "benign neoplasm of skin" and "benign neoplasm of unspecified sites" among women. Although no associations were found for rs7902587 (SLK, STN1) and rs7405776 (HNF1B) variants among women, our analyses among men revealed associations with "disorders of male genital organs" for both variants. Specifically, we found a higher risk of "hyperplasia of the prostate" for the rs7902587 variant and a lower risk of "prostate cancer" and other male genital organ cancers, including "malignant neoplasm of testis", for the rs7405776 variant (Figure 3).

In the phenome-wide analysis using biomarkers and physiological measures as the outcome, we found the strongest associations between the ABO variant and alkaline phosphatase (-0.27 standard deviation (SD), 95% CI -0.028, -0.26), and between the HNF1A-AS1 variant and C-reactive protein (CRP) (-0.12 SD, 95% CI -0.13, -0.11), with the associations slightly stronger among men compared to women (Figure S12). The ABO variant further associated with higher total cholesterol, LDL cholesterol, high density lipoprotein (HDL) cholesterol, apolipoprotein A1 (ApoA1), ApoB, CRP, glycated hemoglobin (HbA1C), glucose, and GGT, and lower triglycerides and aspartate aminotransferase (AST). Interestingly, the ABO variant was associated with lower oestradiol among men while no association was found among women. The rs7953249-G (HNF1A-AS1) allele was associated with higher insulin-like growth factor-1 (IGF-1), total cholesterol, LDL cholesterol, ApoB, cystatin C, and alkaline phosphatase while the same allele associated with lower sex-hormone binding globulin (SHBG), GGT, and urate. The association between rs7953249 and testosterone was directionally different among women (0.01, 95% CI 0.01, 0.02) and men (-0.02, 95% CI -0.03, -0.02). In contrast, rs752590 (PAX8-AS1) - BMI and rs1879586 (PLEKHM1) - IGF-1 associations were found among men but not women. Our PheWAS also identified associations of rs1879586 (PLEKHM1) with seven biomarkers or physiological measures, rs35587371 (MLLT10) with four biomarkers or physiological measures, rs64556822 (GPX6) with two biomarkers, three variants (rs112071820, rs100069690 and rs199661266) each with one biomarker and rs56318008 (WNT4) with one physiological lung measure, in women or men (Figure S12).

**Table S1.** The number of phecodes and summaries of cases across disease category among female participants.

| | | Number of cases with a phecode | | | |
|---|---|---|---|---|---|
| **Disease categories** | **No of phecodes** | **Minimum** | **Median** | **Mean** | **Maximum** |
| Infectious diseases | 20 | 218 | 1007 | 1478 | 5838 |
| Neoplasms | 75 | 210 | 1155 | 2519 | 16600 |
| Endocrine/metabolic | 52 | 200 | 548 | 2344 | 13697 |
| Hematopoietic | 22 | 213 | 534 | 1303 | 6986 |
| Mental disorders | 31 | 211 | 680 | 2597 | 17073 |
| Neurological | 32 | 204 | 530 | 1032 | 7759 |
| Sense organs | 56 | 217 | 546 | 1140 | 12454 |
| Circulatory system | 98 | 207 | 813 | 2452 | 34445 |
| Respiratory | 46 | 221 | 972 | 1757 | 14825 |
| Digestive | 107 | 203 | 1373 | 3148 | 18239 |
| Genitourinary | 96 | 202 | 1230 | 2495 | 15578 |
| Pregnancy complications | 28 | 217 | 1036 | 1315 | 4525 |
| Dermatologic | 50 | 224 | 824 | 1196 | 4094 |
| Musculoskeletal | 72 | 202 | 1287 | 3413 | 28908 |
| Congenital anomalies | 15 | 240 | 393 | 651 | 1255 |
| Symptoms | 21 | 224 | 1400 | 3113 | 23812 |
| Injuries & poisonings | 57 | 208 | 954 | 1825 | 12233 |
| Miscellaneous | 11 | 210 | 2835 | 3421 | 9851 |
| Total | 889 | | | | |

**Table S2.** Ovarian cancer associated variants identified in prior genome-wide association studies (GWAS)

| Genetic variant | Chr. | Nearby gene[1] | Pos. | ea | oa | Phelan et al GWAS: case/control | eaf | Logor | se_logor | p-all[2] | p-min[3] | Discovery GWAS | Included in the study[4] | Each OC subtypes-risk increasing variants[5]: Serous | Endometrioid | Clear Cell | Mucinous |
|---|---|---|---|---|---|---|---|---|---|---|---|---|---|---|---|---|---|
| rs12023270 | 1 | *RSPO1* | 38086578 | T | C | 25509/40941 | 0.259 | 0.0717 | 0.0149 | 1.4E-06 | 4.3E-09 | Kuchenbaecker; Nat Genet (2015) | Yes | | | | |
| rs12131772 | 1 | *AK5* | 77883671 | G | C | 25509/40941 | 0.123 | 0.0324 | 0.0197 | 1.0E-01 | 8.1E-02 | Goode; Nat Genet (2010) | No | | | | |
| rs56318008 | 1 | *WNT4* | 22470407 | T | C | 25509/40941 | 0.146 | 0.0656 | 0.0186 | 4.3E-04 | 4.3E-04 | Kuchenbaecker; Nat Genet (2015) | Yes | | | | |
| rs2165109 | 2 | *MIR4435-2HG, ACOXL* | 111818658 | C | A | 25509/40941 | 0.252 | 0.0751 | 0.0152 | 2.0E-08 | 7.7E-07 | Phelan; Nat Genet (2017) | Yes | | | | |
| rs6755777 | 2 | *HAGLROS, HAGLR, HOXD3* | 177043226 | T | G | 25509/40941 | 0.319 | 0.1063 | 0.0141 | 3.7E-14 | 1.8E-14 | Goode; Nat Genet (2010) | Yes | | | | |
| rs752590 | 2 | *PAX8-AS1* | 113972945 | G | A | 25509/40941 | 0.211 | 0.0264 | 0.0170 | 1.2E-01 | 2.2E-12 | Kelemen; Nat Genet (2015) | Yes | | | | |
| rs112071820 | 3 | *MRPS22* | 138849110 | GCCAGATTCAGAAT | G | 25509/40941 | 0.283 | 0.0279 | 0.0154 | 7.1E-02 | 1.5E-13 | Phelan; Nat Genet (2017) | Yes | | | | |
| rs62274041 | 3 | *TPARP, METTL15P1* | 156435640 | A | G | 25509/40941 | 0.048 | 0.3712 | 0.0289 | 4.5E-38 | 4.0E-49 | Pharoah; Nat Genet (2013) | Yes | | | | |
| rs9870207 | 3 | *CCT6P4, GMNC* | 190525516 | A | G | 25509/40941 | 0.691 | 0.0522 | 0.0145 | 3.2E-04 | 4.5E-08 | Phelan; Nat Genet (2017) | Yes | | | | |
| rs13113999 | 4 | *Y_RNA* | 167187046 | T | G | 25509/40941 | 0.516 | 0.0359 | 0.0141 | 1.1E-02 | 4.7E-08 | Phelan; Nat Genet (2017) | Yes | | | | |
| rs17329882 | 4 | *SYNPO2* | 119949960 | C | A | 25509/40941 | 0.241 | 0.0724 | 0.0154 | 2.5E-06 | 2.5E-06 | Kuchenbaecker; Nat Genet (2015) | Yes | | | | |
| rs4691139 | 4 | *TRIM60, TRIM61* | 165908721 | A | G | 25509/40941 | 0.533 | 0.001926 | 0.01322 | 0.8842 | 1.7E-01 | Couch; PLoS Genet (2013) | No | | | | |
| rs10069690 | 5 | *TERT* | 1279790 | T | C | 25509/40941 | 0.260 | 0.0830 | 0.0150 | 3.4E-08 | 1.5E-12 | Bojesen; Nat Genet (2013) | Yes | | | | |
| rs555025179 | 5 | *MAST4* | 66121089 | GTGACAC | | 25509/40941 | 0.531 | 0.0720 | 0.0136 | 4.5E-08 | 4.5E-08 | Phelan; Nat Genet (2017) | No | | | | |
| rs6456822 | 6 | *GPX6* | 28480635 | T | C | 25509/40941 | 0.691 | 0.0497 | 0.0144 | 5.4E-04 | 3.1E-05 | Kuchenbaecker; Nat Genet (2015) | Yes | | | | |
| rs1400482 | 8 | *LINC00824* | 129541931 | G | A | 25509/40941 | 0.868 | 0.1629 | 0.0203 | 6.9E-16 | 1.5E-24 | Goode; Nat Genet (2010) | Yes | | | | |
| rs150293538 | 8 | *LINC01111* | 77320354 | C | T | 25509/40941 | 0.978 | 0.1330 | 0.0537 | 1.3E-02 | 2.3E-09 | Phelan; Nat Genet (2017) | Yes | | | | |
| rs76837345 | 8 | *CHAMP4C* | 82668818 | G | A | 25509/40941 | 0.066 | 0.1446 | 0.0256 | 1.7E-08 | 9.0E-10 | Pharoah; Nat Genet (2013) | Yes | | | | |
| rs9886651 | 8 | *PVT1* | 128817883 | G | A | 25509/40941 | 0.461 | 0.0583 | 0.0133 | 1.1E-05 | 2.0E-09 | Phelan; Nat Genet (2017) | Yes | | | | |
| chr9:136138765 | 9 | *ABO* | 136138765 | G | GCGCCCACCACTA | 25509/40941 | 0.194 | 0.0994 | 0.0167 | 2.8E-09 | 2.8E-09 | Kuchenbaecker; Nat Genet (2015) | Yes | | | | |
| rs1339552 | 9 | *BNC2* | 16848790 | C | T | 25509/40941 | 0.594 | 0.1287 | 0.0134 | 7.8E-22 | 9.4E-26 | Song; Nat Genet (2009) | Yes | | | | |
| rs320203 | 9 | *RNU6-329P* | 104943226 | C | A | 25509/40941 | 0.850 | 0.0118 | 0.0187 | 5.3E-01 | 1.7E-08 | Phelan; Nat Genet (2017) | Yes | | | | |
| rs35587371 | 10 | *MLLT10* | 21878831 | A | T | 25509/40941 | 0.309 | 0.0916 | 0.0139 | 4.0E-11 | 5.2E-10 | Pharoah; Nat Genet (2013) | Yes | | | | |
| rs7902587 | 10 | *SLK, STN1* | 105694301 | T | C | 25509/40941 | 0.098 | 0.0660 | 0.0224 | 3.2E-03 | 4.0E-08 | Phelan; Nat Genet (2017) | Yes | | | | |
| rs7953249 | 12 | *HNF1A-AS1* | 120965921 | G | A | 25509/40941 | 0.418 | 0.0801 | 0.0131 | 5.0E-10 | 5.0E-10 | Phelan; Nat Genet (2017) | Yes | | | | |
| rs8044477 | 16 | *Metazoa_SRP* | 58976591 | G | A | 25509/40941 | 0.623 | 0.0297 | 0.0137 | 3.1E-02 | 2.4E-03 | Kuchenbaecker; Nat Genet (2015) | Yes | | | | |
| rs12938171 | 17 | *BRIP1* | 59980355 | A | G | 25509/40941 | 0.014 | 0.0284 | 0.0641 | 6.6E-01 | 6.0E-01 | Rafnar; Nat Genet (2011) | No | | | | |
| rs1879586 | 17 | *PLEKHM1* | 43567337 | G | C | 25509/40941 | 0.181 | 0.1257 | 0.0171 | 1.8E-13 | 1.8E-13 | Couch; PLoS Genet (2013) | Yes | | | | |
| rs199661266 | 17 | *ATAD5* | 29181220 | A | AT | 25509/40941 | 0.720 | 0.0788 | 0.0149 | 1.3E-07 | 1.3E-07 | Kuchenbaecker; Nat Genet (2015) | Yes | | | | |
| rs7207826 | 17 | *SKAP1* | 46500673 | C | T | 25509/40941 | 0.268 | 0.1048 | 0.0146 | 7.7E-13 | 1.2E-14 | Pharoah; Nat Genet (2013) | Yes | | | | |
| rs7405776 | 17 | *HNF1B* | 36093022 | A | G | 25509/40941 | 0.375 | 0.0508 | 0.0136 | 1.9E-04 | 1.9E-10 | Pharoah; Nat Genet (2013) | Yes | | | | |
| rs8098244 | 18 | *LAMA3* | 21405553 | A | G | 25509/40941 | 0.275 | 0.0382 | 0.0149 | 1.0E-02 | 3.8E-08 | Phelan; Nat Genet (2017) | Yes | | | | |
| rs4808075 | 19 | *BABAM1 ,USHBP1* | 17390291 | C | T | 25509/40941 | 0.296 | 0.1224 | 0.0144 | 1.5E-17 | 3.2E-26 | Bolton; Nat Genet (2010) | Yes | | | | |
| rs688187 | 19 | *IFNL3P1, IFNL3* | 39732752 | A | G | 25509/40941 | 0.313 | 0.0108 | 0.0154 | 4.8E-01 | 1.2E-22 | Kelemen; Nat Genet (2015) | Yes | | | | |
| rs6005807 | 22 | *TTC28* | 28934313 | C | T | 25509/40941 | 0.899 | 0.1144 | 0.0223 | 2.9E-07 | 4.5E-09 | Phelan; Nat Genet (2017) | Yes | | | | |

rs555025179 (merged into rs56076405) - leading and proxy SNPs ($r^2$>0.8) not available in the UK Biobank imputed genome wide data.

[1] nearby gene from GWAS catalog

[2] p-all is p-value for association with overall OC in the Phelan et al GWAS

[3] p-min is the smallest p-value for association with overall or subtypes of OC in the Phelan et al GWAS

[4] variants that were genome-wide significant in the Phelan et al GWAS or those who were identified in the prior GWASs and were significant at p-value of 0.05 (see p-min column) in the Phelan et al GWAS were included for the analysis.

[5] list of variants associated with each subtype of OC at genome-wide significant threshold (dark green) or at p<0.05 (light green) in in Phelan et al GWAS

**Table S3**. Functional blocks based on the PhenoScanner search for diseases/traits associated with ovarian cancer risk variants (or proxy at $R^2 \geq 0.8$) at genome-wide significant threshold ($p \leq 5 \times 10^{-8}$).

| Functional blocks | Chr. | Nearby gene | Genetic variant | Traits |
|---|---|---|---|---|
| **Cancers (not including OC)** | 5 | *TERT* | rs10069690 | Breast cancer, Glioma, Glioblastoma, Non-glioblastoma glioma, Testicular germ cell tumor testicular cancer, Prostate cancer, Thyroid cancer |
| | 10 | *SLK/STN1* | rs7902587 | Thyroid cancer |
| | 17 | *HNF1B* | rs7405776 | Malignant neoplasm of prostate, Self-reported prostate cancer |
| | 19 | *BABAM1, USHBP1* | rs4808075 | Breast cancer, Cancer pleiotropy |
| **Hormonal and gynaecological problems** | 1 | *WNT4* | rs56318008 | Endometriosis, Female genital prolapse, Self-reported uterine fibroids |
| | 8 | *LINC00824* | rs1400482 | Self-reported hypothyroidism or myxoedema |
| | 17 | *PLEKHM1* | rs1879586 | Hair or balding pattern: pattern 3, Hair or balding pattern: pattern 4, Relative age of first facial hair, Relative age voice broke |
| | 17 | *HNF1B* | rs7405776 | Endometriosis, Female genital prolapse, Self-reported uterine fibroids |
| **Anthropometrics** | 1 | *WNT4* | rs56318008 | Sitting height, Lean body mass, Impedance of leg right |
| | 2 | *HAGLROS, HAGLR* | rs6755777 | Impedance of arm left |
| | 3 | *METTL15P1, LINC00886* | rs62274041 | Height |
| | 10 | *MLLT10* | rs35587371 | Body fat percentage, Body mass index, Hip circumference, Waist circumference, Weight, Body fat mass |
| | 10 | *SLK/STN1* | rs7902587 | Height |
| | 17 | *PLEKHM1* | rs1879586 | Height, Comparative height size at age 10, Impedance measures |
| **Cardiometabolic** | 12 | *HNF1A-AS1* | rs7953249 | Total cholesterol, LDL cholesterol, Self-reported high cholesterol, Treatment with cholesterol lowering medication, Inflammation, Coronary artery disease |
| | 17 | *HNF1B* | rs7405776 | Diabetes diagnosed by doctor, Self-reported diabetes, Treatment with metformin |
| **Biomarkers** | 9 | *ABO* | chr9:136138764-136138765 | Alkaline phosphatase |
| | 10 | *MLLT10* | rs35587371 | Sodium in urine |
| | 12 | *HNF1A-AS1* | rs7953249 | Serum gamma glutamyl transferase activity in adults, C-reactive protein, Plasma C reactive protein female |
| **Blood counts** | 2 | *MIR4435-2HG, ACOXL* | rs2165109 | Granulocyte percentage of myeloid white cells, Mean corpuscular hemoglobin, Mean corpuscular volume, Monocyte percentage of white cells, Red blood cell count |
| | 2 | *PAX8-AS1* | rs752590 | Hematocrit, Hemoglobin concentration, Red blood cell count |
| | 5 | *TERT* | rs10069690 | Mean corpuscular hemoglobin, Plateletcrit, Red blood cell count |
| | 6 | *GPX6* | rs6456822 | Mean corpuscular hemoglobin, Red cell distribution width, Reticulocyte count, Reticulocyte fraction of red cells |
| | 9 | *ABO* | chr9:136138764-136138765 | Sum basophil neutrophil counts, Sum eosinophil basophil counts, Granulocyte count, Hematocrit, Hemoglobin concentration, Red blood cell count, Red cell distribution width, White blood cell count, Lymphocyte percentage of white cells, Monocyte count, Myeloid white cell count, Platelet distribution width |
| | 9 | *BNC2* | rs1339552 | High light scatter percentage of red cells, High light scatter reticulocyte count |
| | 12 | *HNF1A-AS1* | rs7953249 | Platelet count, Plateletcrit |
| | 17 | *PLEKHM1* | rs1879586 | Eosinophil count, Eosinophil percentage of granulocytes, Eosinophil percentage of white cells, Hematocrit, Hemoglobin concentration, Mean platelet volume, Neutrophil percentage of granulocytes, Neutrophil percentage of white cells, Red blood cell count, Red cell distribution width, Reticulocyte fraction of red cells, Sum eosinophil basophil counts. |
| | 17 | *SKAP1* | rs7207826 | Mean corpuscular hemoglobin concentration, Red cell distribution width, Reticulocyte count, Reticulocyte fraction of red cells |
| **Respiratory** | 1 | *WNT4* | rs56318008 | Forced expiratory volume in 1-second, Forced vital capacity |
| | 3 | *METTL15P1, LINC00886* | rs62274041 | Forced vital capacity |
| | 5 | *TERT* | rs10069690 | Fibrotic idiopathic interstitial pneumonias pulmonary fibrosis |
| | 10 | *MLLT10* | rs35587371 | Forced vital capacity |
| | 17 | *PLEKHM1* | rs1879586 | Forced expiratory volume in 1-second, Forced vital capacity, Peak expiratory flow |
| **Miscellaneous** | 1 | *WNT4* | rs56318008 | Total body bone mineral density, Heel bone mineral density, Gestational age at birth maternal effect |
| | 6 | *GPX6* | rs6456822 | Disorders of mineral metabolism, Self-reported malabsorption or coeliac disease |
| | 9 | *ABO* | chr9:136138764-136138765 | Ferritin |
| | 9 | *BNC2* | rs1339552 | Melanocytic naevi, Skin freckles, Melanosis |
| | 10 | *MLLT10* | rs35587371 | Usual walking pace |
| | 12 | *HNF1A-AS1* | rs7953249 | Antennary fucosylated glycans, Desialylated Glycan Peak 7, Desialylated Glycan Peak 9, N glycan levels, Polysaccharides |
| | 17 | *PLEKHM1* | rs1879586 | Hand grip strength, Heel bone mineral density, Treatment with paracetamol, "Pain type experienced in last month: headache", "Qualifications: college or university degree", Getting up in morning, Nap during day, Daytime dozing or sleeping, Mood swings, Frequency of tenseness or restlessness in last 2 weeks, Neuroticism score, Sensitivity or hurt feelings |
| | 19 | *IFNL3P1* | rs688187 | Anti-hepatitis C virus treatment response Null virologic responder to pegylated interferon alpha and ribavarin therapy |

**Table S4:** Descriptions of covariates included in the analyses*

| Covariate | Categorisation or description [as used] | UK Biobank field ID |
|---|---|---|
| **Age** | Age in years used as continous variable | 21022 [Age at recruitment] |
| **Assessment centre** | | 54 [UK Biobank Assessment Centres] |
| **Education** | Intermediate (NVQ/CSE/A-levels) or none<br>High (degree/professional) | 6138 and 10722 (pilot) [“Which of the following qualifications do you have?”] |
| **Employment status** | Unemployed<br>Retired<br>Lowest working hour (first quartile)<br>Second quartile<br>Third quartile<br>Highest working hour (fourth quartile) | 6142 and 20119 (pilot) [Which of the following describes your current (“working”) situation?<br>767 [In a typical WEEK, how many hours do you spend at work? (Do not include hours travelling to and from work" |
| **Townsend deprivation index (TDI)** | Low (higher socioeconomic status) Vs High deprivation index (lower socioeconomic status) using median | 189 [Townsend deprivation index calculated immediately prior to participating to participant joining the UK Biobank, based on the preceding national census output areas] |
| **Alcohol consumption** | Never<br>Previous<br>Previous | 20117 ["Alcohol drinker status"] |
| **Regular physical activity** | MET/week, constructed using the listed variables | 884 [In a typical WEEK, on how many days did you do 10 minutes or more of moderate physical activities like carrying light loads, cycling at a normal pace? (Do not include walking)]<br>894 [How many minutes did you usually spend doing moderate activities on a typical DAY?]<br><br>904 [In a typical WEEK, how many days did you do 10 minutes or more of vigorous physical activity? (These are activities that make you sweat or breathe hard such as fast cycling, aerobics, heavy lifting)]<br>914 ["How many minutes did you usually spend doing vigorous activities on a typical DAY?"] |
| **Body mass index** | Weight/height square ($kg/m^2$) | 21002 [Measured weight]<br>50 [Standing height measure using a Seca 202 device] |

* Information used were those collected at recruitment/baseline visits. NVQ, National Vocational Qualification; CSE, Certificate of Secondary Education; A-levels, Advanced-level

**Table S5.** The association between each potential confounding factors and ovarian cancer risk score.

| | **Ovarian cancer (OC)-GRS** | | **Serous OC-GRS** | | **Endometrioid OC-GRS** | | **Clear cell OC-GRS** | | **Mucinous OC-GRS** | |
|---|---|---|---|---|---|---|---|---|---|---|
| | OR (95% CI) | p | OR (95% CI) | p | OR (95% CI) | p | OR (95% CI) | p | OR (95% CI) | p |
| Age | 1.00 (1.00,1.00) | 0.99 | 1.00 (1.00,1.01) | 0.20 | 1.00 (0.99,1.01) | 0.51 | 1.00 (1.00,1.01) | 0.22 | 0.99 (0.99,1) | 0.02 |
| Education | 1.00 (1.00,1.00) | 0.76 | 1.00 (1.00,1.01) | 0.74 | 0.99 (0.98,1.00) | 0.04 | 1.00 (0.99,1.01) | 0.95 | 1.00 (1.00,1.01) | 0.53 |
| Townsend deprivation index (TDI) | 1.00 (1.00,1.00) | 0.92 | 1.00 (1.00,1.01) | 0.58 | 1.01 (1.00,1.01) | 0.20 | 1.00 (0.99,1.01) | 0.94 | 1.01 (1.00,1.01) | 0.02 |
| Body mass index (BMI) | 1.00 (1.00,1.01) | 0.08 | 1.00 (1.00,1.01) | 0.49 | 1.01 (1.00,1.02) | 0.11 | 1.00 (0.99,1.01) | 0.75 | 1.00 (0.99,1.00) | 0.37 |
| Physical activity | 1.00 (0.99,1.00) | 0.03 | 0.99 (0.99,1.00) | 0.02 | 1.00 (0.99,1.00) | 0.39 | 0.99 (0.99,1.00) | 0.18 | 1.00 (0.99,1.00) | 0.09 |
| Alcohol consumption | 1.00 (1.00,1.01) | 0.39 | 1.00 (1.00,1.01) | 0.54 | 1.00 (0.99,1.02) | 0.70 | 1.00 (0.99,1.01) | 0.95 | 1.00 (0.99,1.01) | 0.82 |

Effect estimates are from logistic regression regression in analyses restricted to women. Age coded as binary (below 60 years vs 60 years age and above), education coded as binary (College or University degree vs. other levels including none), Townsend deprivation index (less deprived vs more deprived; using a median), body mass index codes as binary (below 30 kg/m3 vs 30 $kg/m^2$), physical activity codes as binary(high versus low, based on the median MET/week), and alcohol consumption (never vs. ex- or current drinker). All analyses adjusted for assessment centre (as n-1 dummy variable), birth location (as n-1 dummy variable), types of SNP array, 40 principal components. None were associated with the risk score at Bonferroni corrected threshold of (0.05/6=0.008).

**Table S6.** Genome-wide significant variant – diseases associations identified in the PheWAS, and confirmation of novelty using the reported associations in the GWAS Catalog.

| Genetic variant [nearby gene] | Phecode | Descriptions | Categories | Population | $p_{phewas}$ | Is it novel? [if no, PubMed ID] | Remarks |
|---|---|---|---|---|---|---|---|
| **rs56318008 [*WNT4*]** | 618 | Female genitals prolapse | Genitourinary | Women | 1.1E-11 | No [34594039, 32184442] | rs3820282 in LD with rs56318008 [*WNT4*] ($r^2$=0.904) associated with uterine prolapse and pelvic organ prolapse in European ancestry GWAS. |
| **rs10069690 [*TERT*]** | 218 | Benign neoplasm of uterus | Neoplasms | Women | 7.5E-15 | No [30194396, 31649266] | The variant was previously identified for benign neoplasm of the uterus. |
| | 218.1 | Uterine leiomyoma | Neoplasms | Women | 8.4E-15 | No [30194396, 31649266] | |
| | 702.2 | Seborrheic keratosis | Dermatologic | Women | 1.6E-08 | Yes? | |
| | | | | Men | 6.1E-07 | | |
| | 185 | Cancer of prostate | Neoplasms | Men | 2.0E-09 | No [25939597,29892016, 33398198, 32887889, 33293427, 21743467, 31562322] | Two SNPs (rs2242652 and rs7725218) in LD with rs10069690 [*TERT*] ($r^2$ ≥0.599) associated with prostate cancer in European ancestry population. |
| **rs6456822 [*GPX6*]** | 275 | Disorders of mineral metabolism | Endocrine/metabolic | Women | 5.5E-04 | | Association with related traits (serum glycated hemoglobin levels) were found for LD variant (rs34979126, $r^2$ =0.224) in trans-ancestral GWAS [34059833]. A link between iron deficiency anemia and higher glycated hemoglobin found in prior study [PubMed ID: 35000894] |
| | | | | Men | 9.4E-14 | Yes? | |
| | 275.1 | Disorders of iron metabolism | Hematopoietic | Women | 5.9E-21 | Yes? | |
| | | | | Men | 4.5E-42 | Yes? | |
| | 557.1 | Celiac disease | Digestive | Women | 4.5E-08 | Yes? | |
| | | | | Men | 0.15 | | |
| **rs1339552 [*BNC2*]** | 216 | Benign neoplasm of skin | Neoplasms | Women | 1.5E-10 | No [32041948, 30166351, 33549134, 34290314, 25705849, 27539887, 27424798, 31174203, 31174203, 34594039] | Five variants in LD with rs1339552 [*BNC2*] (r2>0.841) associated with non-melanoma skin cancer and skin pigmentation. |
| | | | | Men | 7.1E-05 | | |
| | 229 | Benign neoplasm of unspecified sites | Neoplasms | Women | 1.3E-09 | Yes? | |
| | | | | Men | 5.1E-04 | | |
| **Chr9:136138765 [*ABO*]** | 415 | Pulmonary heart disease | Circulatory system | Women | 2.8E-12 | ? | ABO locus is very pleiotropic loci with association reported for multiple cardiovascular and cerebrovascular diseases, and related biomarkers. For instance, 15 LD variants ($r^2$ ≥0.4) were associated with different cardiovascular diseases including heart failure, coronary heart diseases, stroke, thromboembolism, and antithrombotic agents. However, the locus was not previously reported for the diseases mentioned in our study. |
| | | | | Men | 4.6E-20 | ? | |
| | 451 | Phlebitis and thrombophlebitis | Circulatory system | Women | 1.1E-17 | ? | |
| | | | | Men | 1.3E-31 | ? | |
| | 451.2 | Phlebitis of lower extremities* | Circulatory system | Women | 4.1E-17 | ? | |
| | | | | Men | 1.2E-33 | ? | |
| | 459 | Other disorders of circulatory system | Circulatory system | Women | 1.3E-18 | ? | |
| | | | | Men | 4.9E-13 | ? | |
| | 459.9 | Circulatory disease NEC | Circulatory system | Women | 6.7E-19 | ? | |
| | | | | Men | 2.5E-13 | ? | |
| **rs7902587 [*SLK, STN1*]** | 600 | Hyperplasia of prostate | Genitourinary | Men | 1.0E-09 | Yes? | |
| **rs7405776 [*HNF1B*]** | 185 | Cancer of prostate | Neoplasms | Men | 2.0E-21 | No [26 studies] | Nine variants in LD with rs7405776 [*HNF1B*] (r2>0.209) were identified for prostate cancer or prostate-specific antigen levels or testicular germ cell tumor at genome-wide significant level. Reported in previous 26 previous studies. |
| | 187 | Cancer of other male genital organs | Neoplasms | Men | 1.0E-12 | No [26 studies] | |
| | 187.1 | Neoplasm of unspecified male genital organ | Neoplasms | Men | 7.0E-13 | No [26 studies] | |
| | 187.2 | Malignant neoplasm of testis | Neoplasms | Men | 1.0E-12 | No [26 studies] | |

Association estimates are from PheWAS with logistic regression model adjusting for age, assessment centre, birth location, type of genotyping array and 40 principal components.

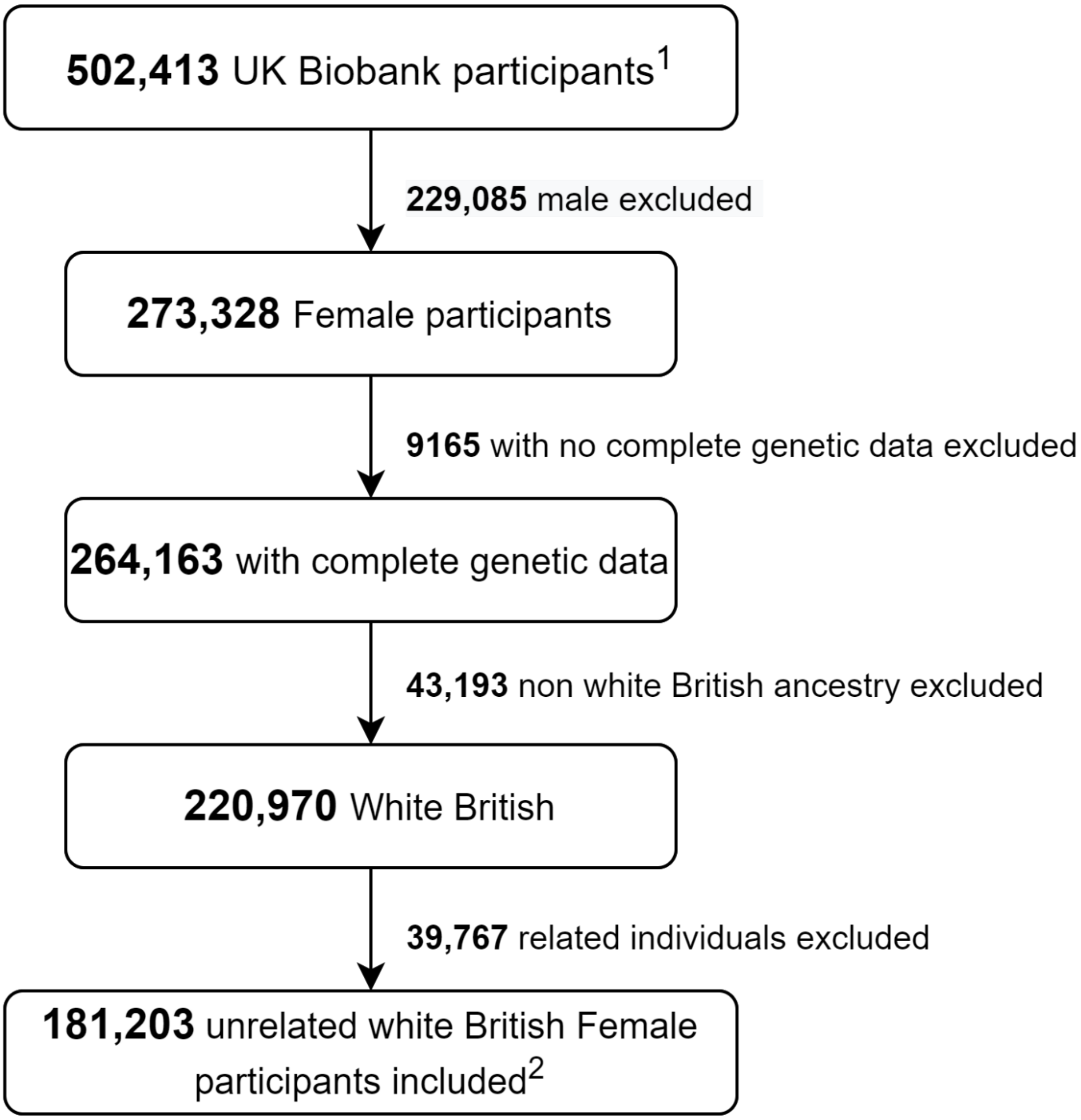


**Figure S1.** Study sample flow for the MR-PheWAS analysis

[1] Actively consented participant
[2] actual final sample size for each outcome are slightly different as individuals with diagnosis from the same group were excluded from the control.

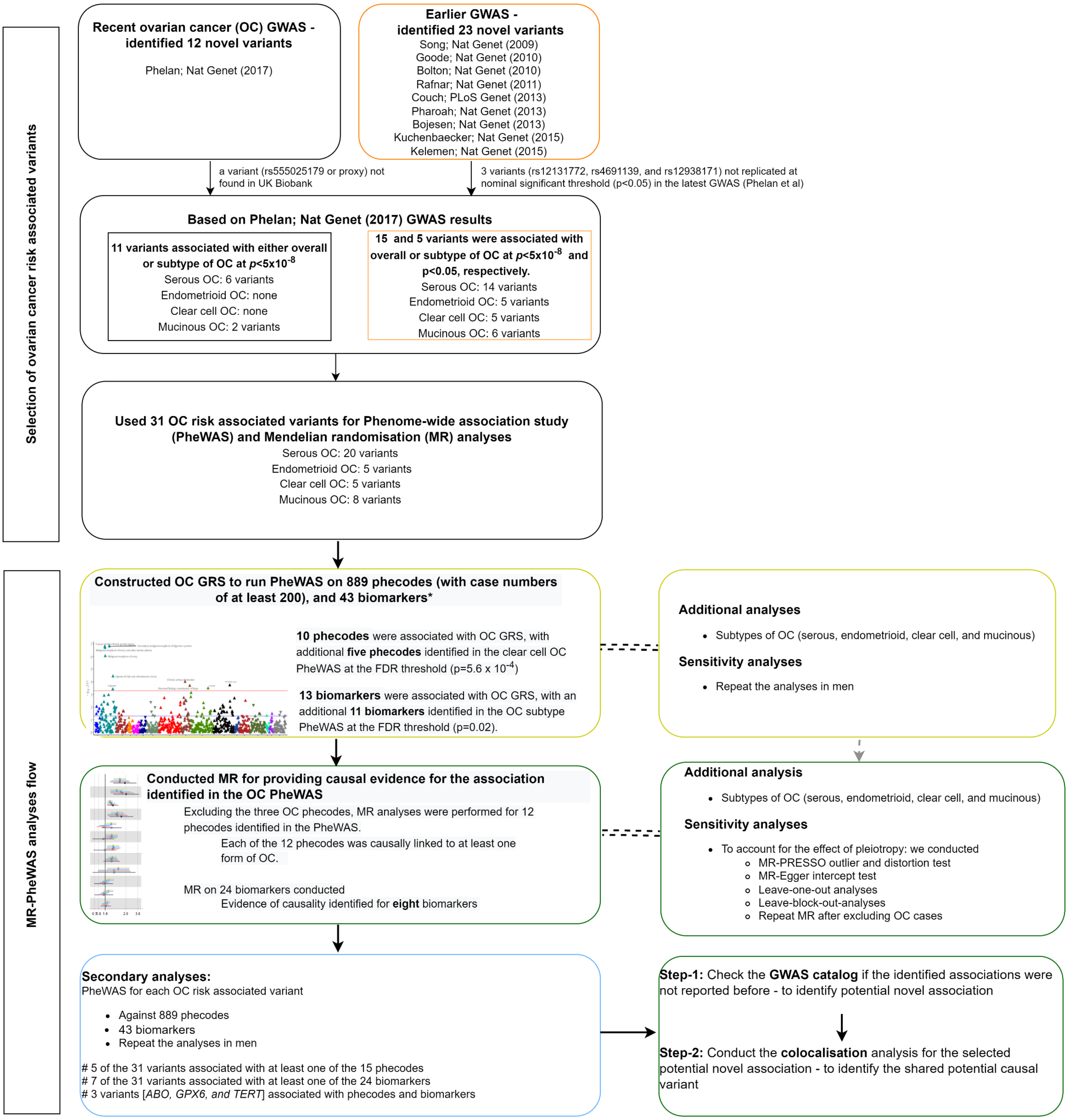


**Figure S2**. Genetic variant selection and analysis strategy.

* The label biomarkers are here used to denote 43 outcomes that include 30 serum and four urine biomarkers, nine physiological measures.

**Panel A**

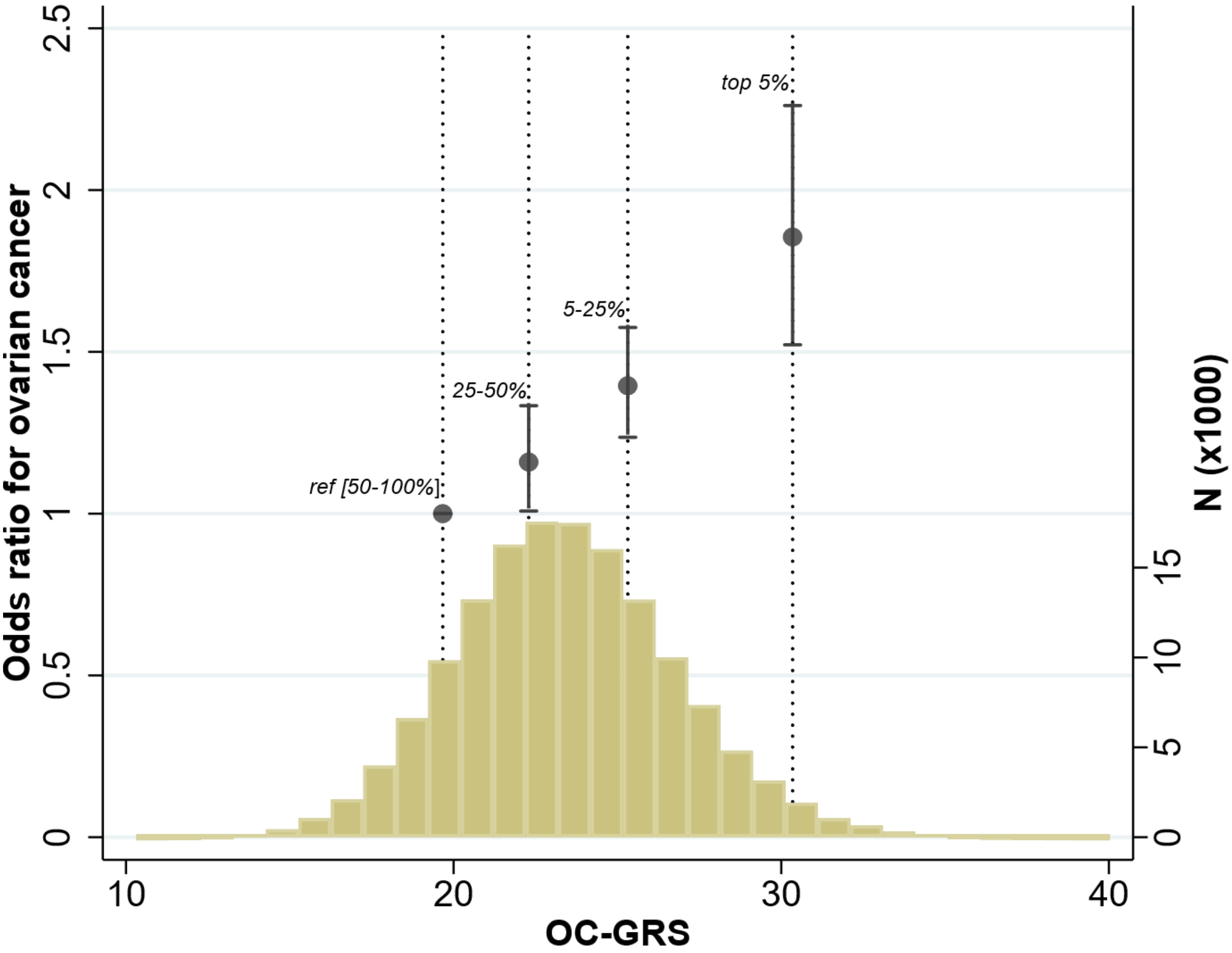


| GRS | Oucomes | Cases,n | Controls,n | OR (95%CI) | p |
|---|---|---|---|---|---|
| Ovarian cancer (OC)-GRS | OC-phecode | 2072 | 122255 | 1.03 (1.02, 1.05) | 1.0E-06 |
| | OC-registry | 1863 | 145583 | 1.06 (1.04,1.07) | 4.7E-16 |
| Serous OC-GRS | Serous OC | 588 | 145583 | 1.12 (1.08, 1.16) | 2.2E-09 |
| Endometrioid OC GRS | Endometrioid OC | 119 | 145583 | 1.24 (1.07,1.44) | 4.2E-03 |
| Clear Cell OC GRS | Clear Cell OC | 75 | 145583 | 1.06 (0.91,1.25) | 4.4E-01 |
| Mucinous OC GRS | Mucinous OC | 80 | 145583 | 1.01 (0.90,1.13) | 8.9E-01 |

**Figure S3.** Distribution of ovarian genetic risk score (GRS) and the association with ovarian cancer in UK Biobank (**Panel A**) and the association between each GRS and its corresponding subtype of ovarian cancer (**Panel B**). In Panel A, the y axis in the left shows the odds ratio for OC for each genetic risk group divided as top 5% (highest genetic risk group, 5-25%, 25-50% and 50-100% (lowest genetic risk group, used as reference). The right y axis shows the distribution of GRS where N reflects the frequency in the scale of thousands. In Panel B (the table), the odds ratios are shown per one risk allele higher. OC-phecode: is ovarian cancer cases defined based Phecode mapping algorithms. OC-registry: is ovarian cancer defined based on the information provided from hospital inpatient and cancer registry data (see supplementary methods). P (in Panel B) – is the p-value from logistic regression of ovarian cancer or the subtypes against the respective GRS after adjusting for age, centre, 40 PC and genotyping array. The F-statistic for the association between OC-GRS and ovarian cancer were 23.7, which this value generated from regression model.

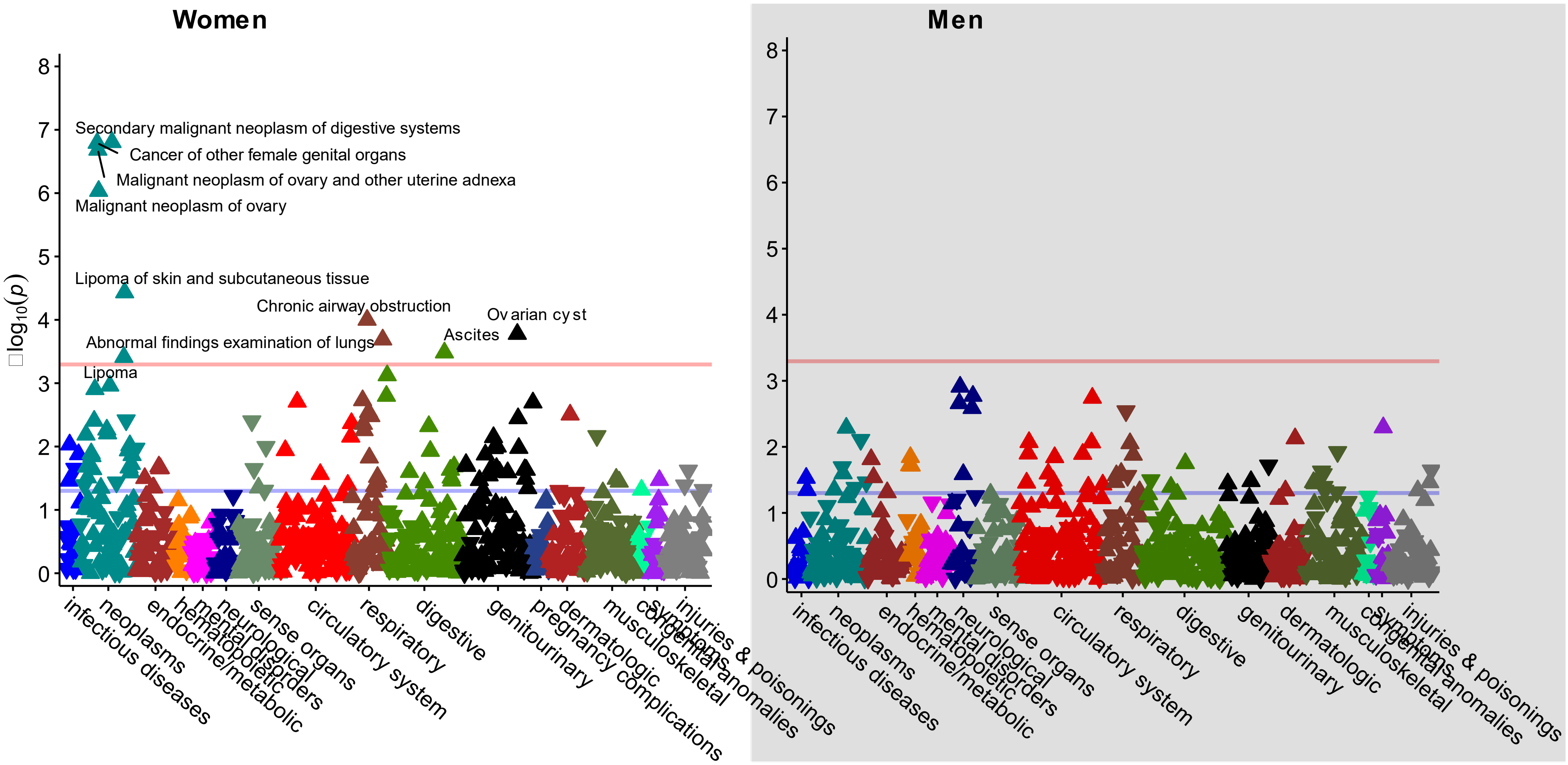


**Figure S4**. Manhattan plots for phenome-wide analysis of ovarian cancer genetic risk score conducted in women (left) and men (right).

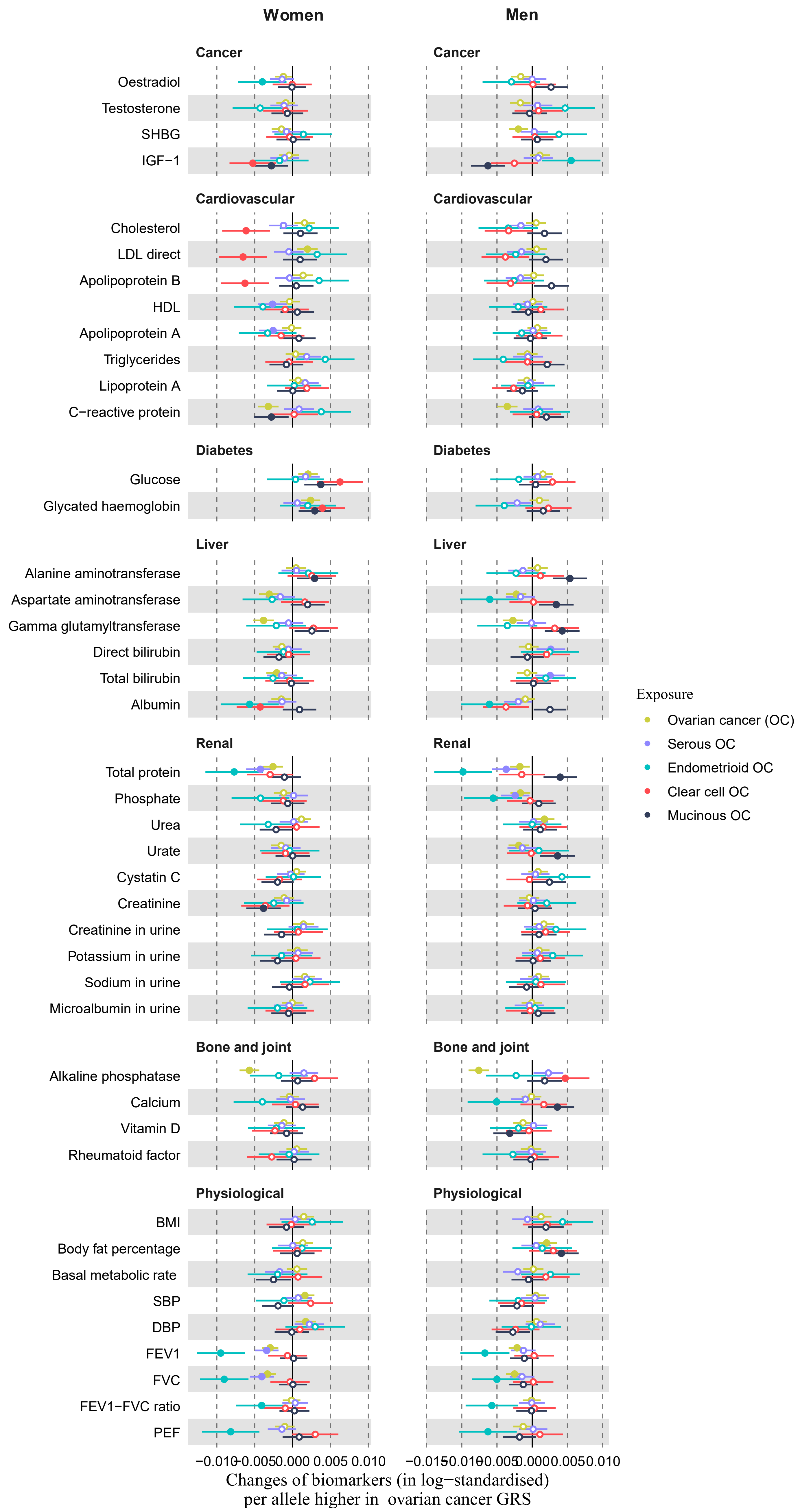


**Figure S5**. Phenome-wide analysis of ovarian cancer genetic risk score against serum and urine biomarkers and physiological measures in women (left) and men (right).

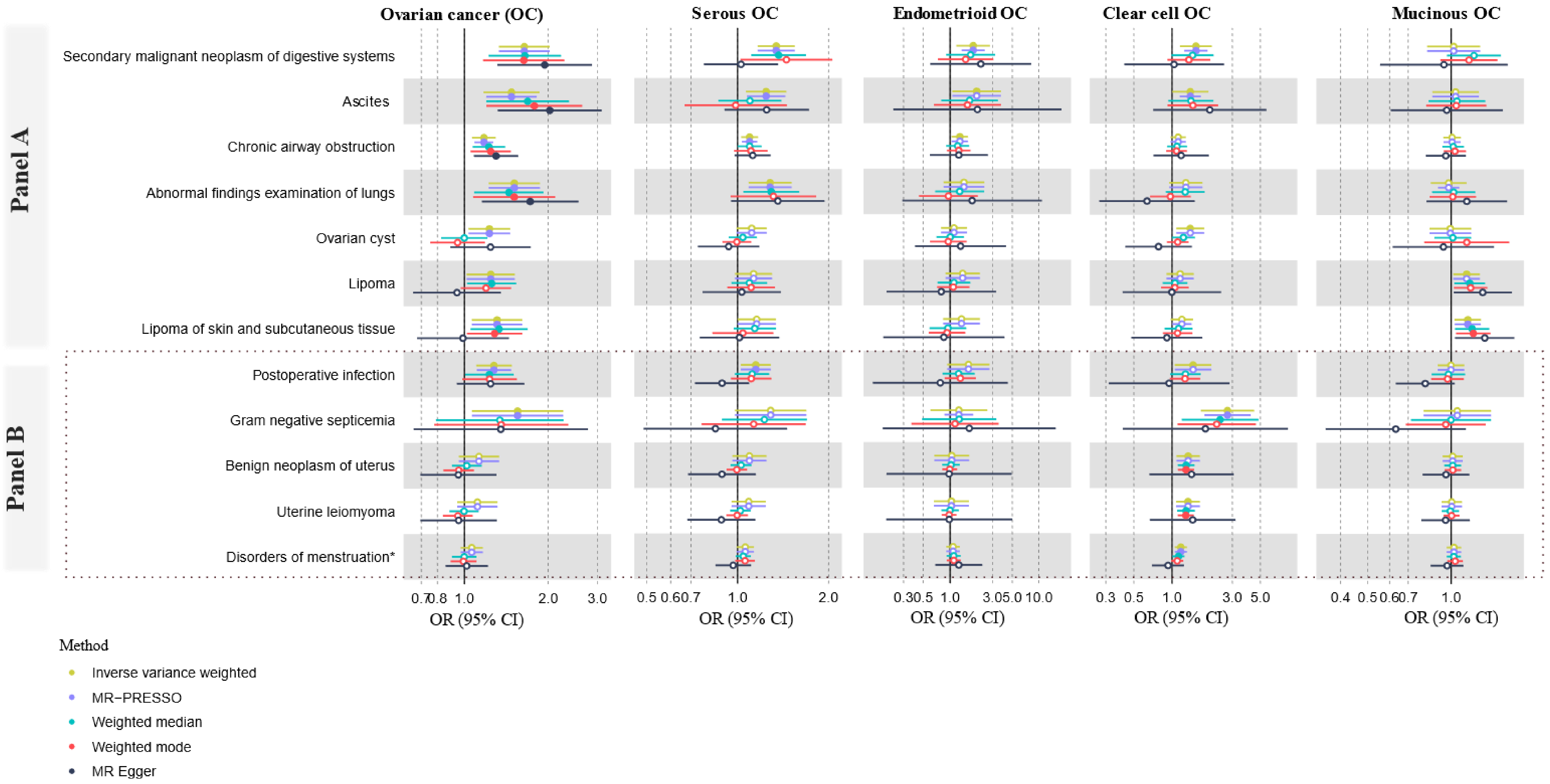


**Figure S6.** Mendelian randomisation estimates from different methods for different types of ovarian cancer. The disease outcomes under **Panel A** are identified in the overall OC PheWAS, and those diseases under **Panel B** were identified only in the clear cell carcinoma PheWAS. MR-Egger intercept test $P_{intercept}$ = 0.01 for serous OC-postoperative infection association; no evidence for pleiotropy identified for other comparisons ($P_{intercept} \geq 0.05$).

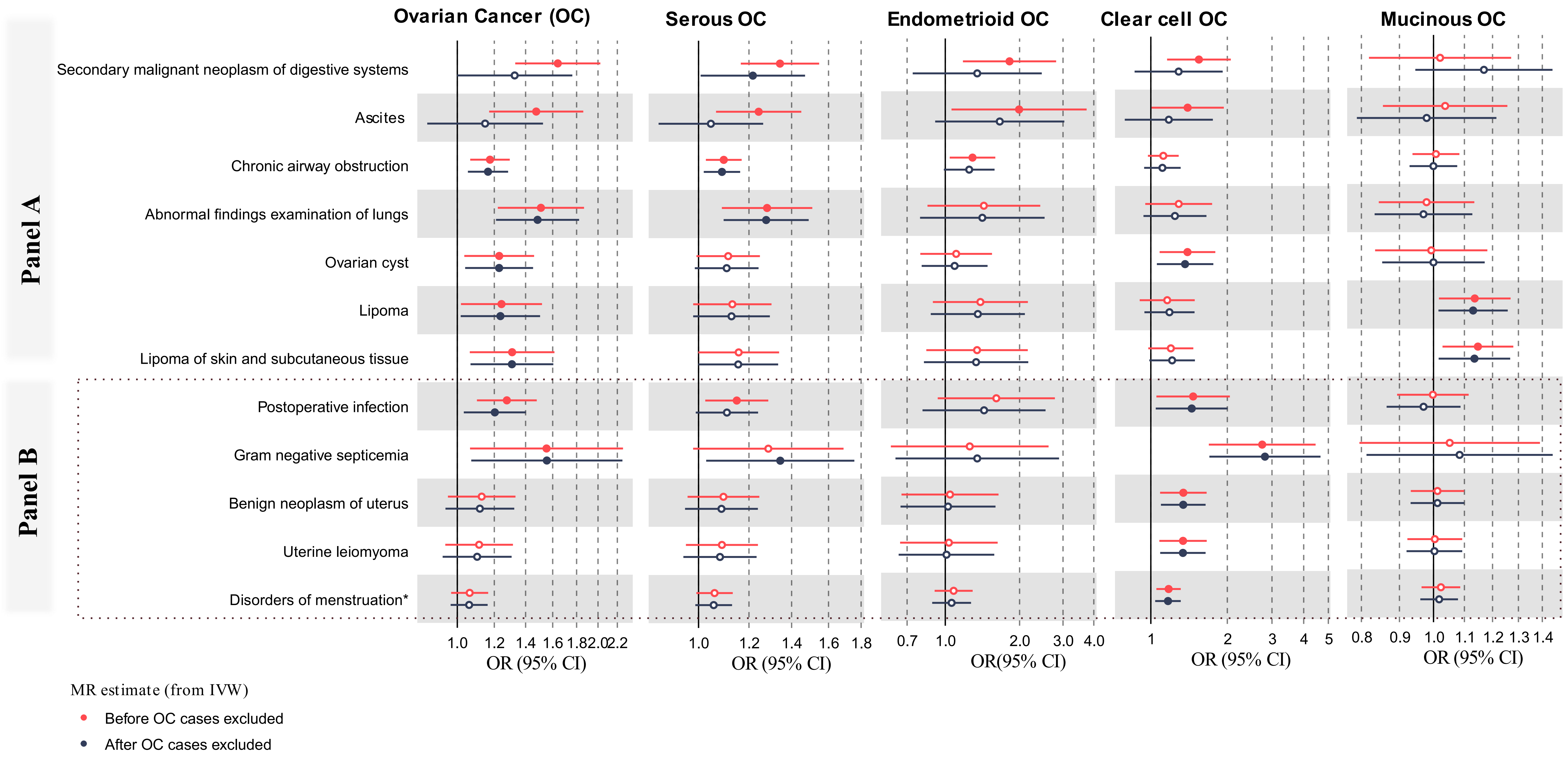


**Figure S7.** Mendelian randomisation estimates before (main analysis) and after (sensitivity analysis) removing ovarian cancer cases. The disease outcomes under **Panel A** are identified in the overall OC PheWAS, and those diseases under **Panel B** were identified only in the clear cell carcinoma PheWAS.

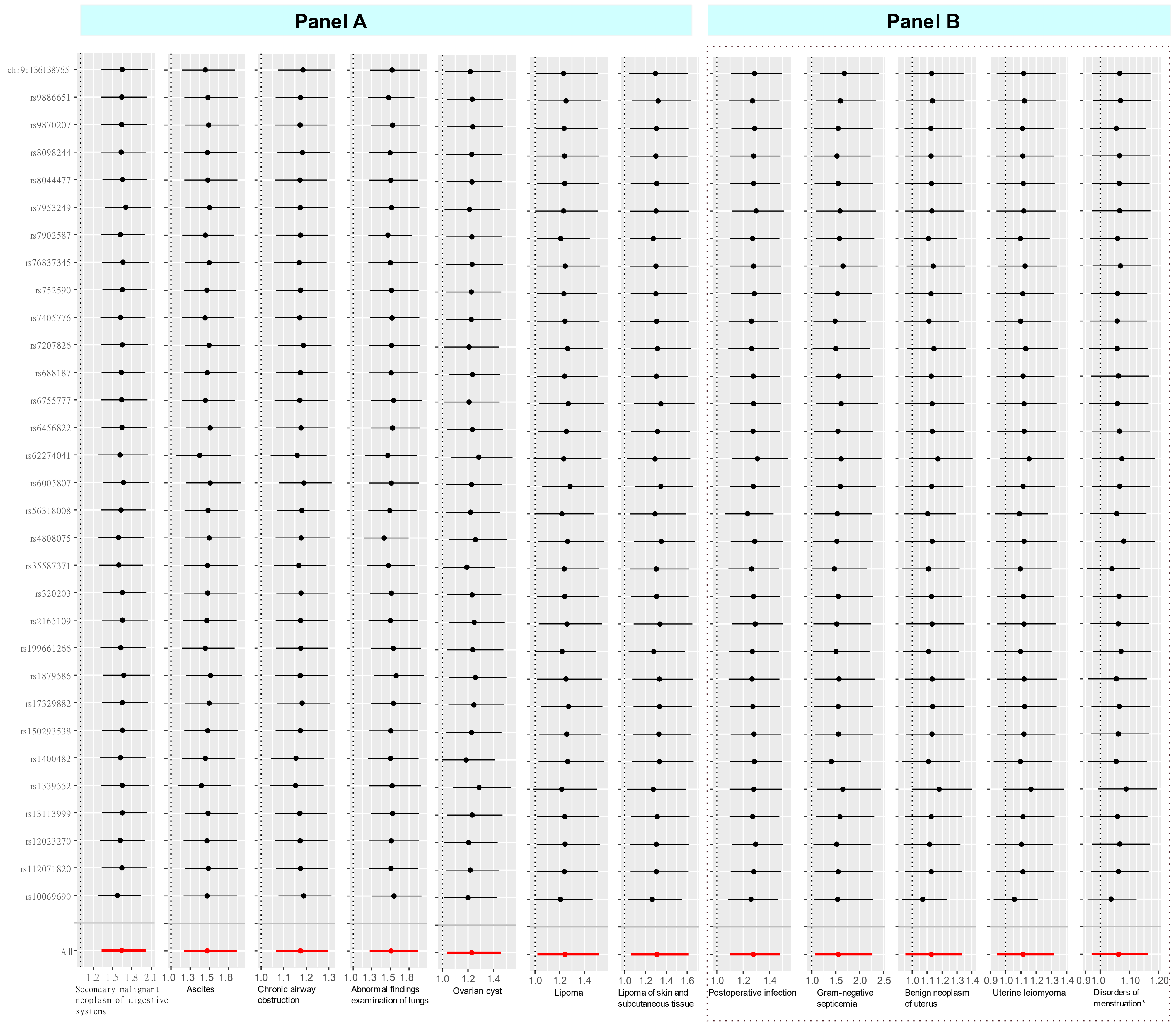


**Figure S8.** **L**eave-one-out mendelian randomisation analyses of ovarian cancer and 12 diseases outcomes. The disease outcomes under **Panel A** are identified in the overall OC PheWAS, and those diseases under **Panel B** were identified only in the clear cell carcinoma PheWAS. Effect estimates shown are OR (95% CI) from the inverse variance weight method.

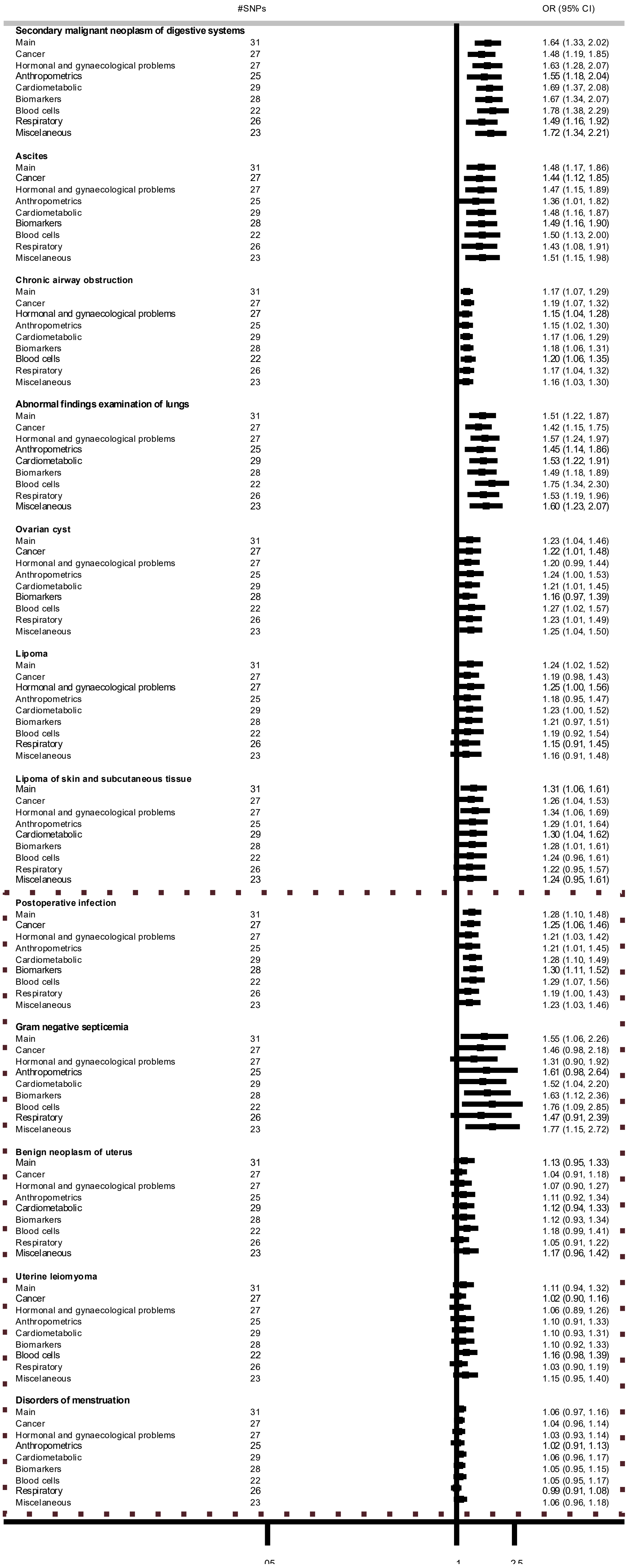


**Figure S9.** Mendelian randomisation estimates from leave-block analyses of ovarian cancer and selected disease outcomes. Estimates in the dotted box shows the genetic OC associations with diseases that were identified in the phenome-wide analyses of clear cell OC. No additional signals were detected in phenome-wide analyses of other OC subtypes.

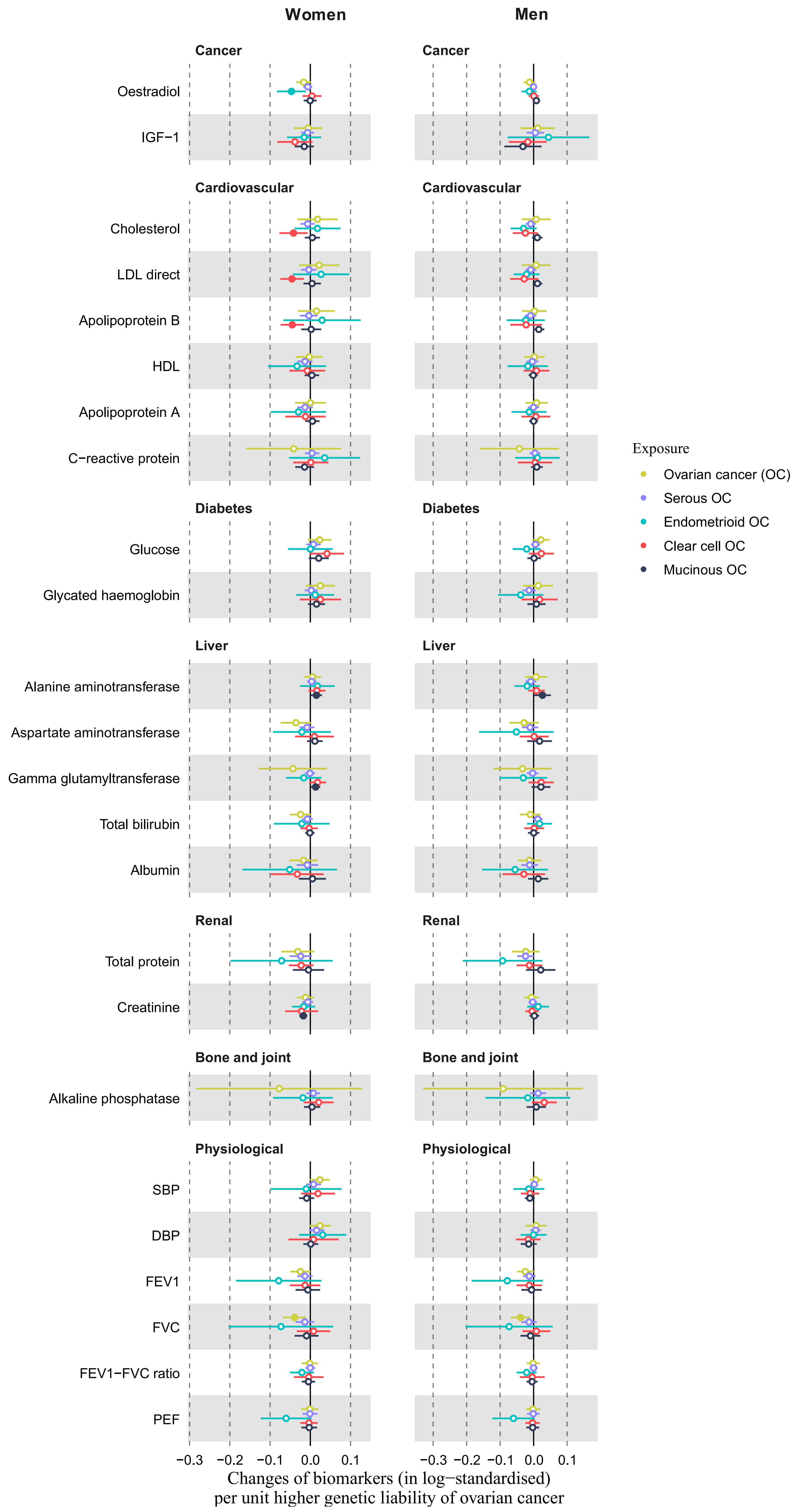


**Figure S10**. Mendelian randomisation estimates of subtypes of ovarian cancer with serum and blood biomarkers, and physiological measures among women (left panel) and men (right panel). Estimates in solid dot are significant at p ≤0.05 threshold. $P_{intercept}$ ≥0.11 for all, and no outlier detect from leave-one-out analyses (results not shown). All biomarkers were log-transformed and standardised to mean zero and standard deviation of one.

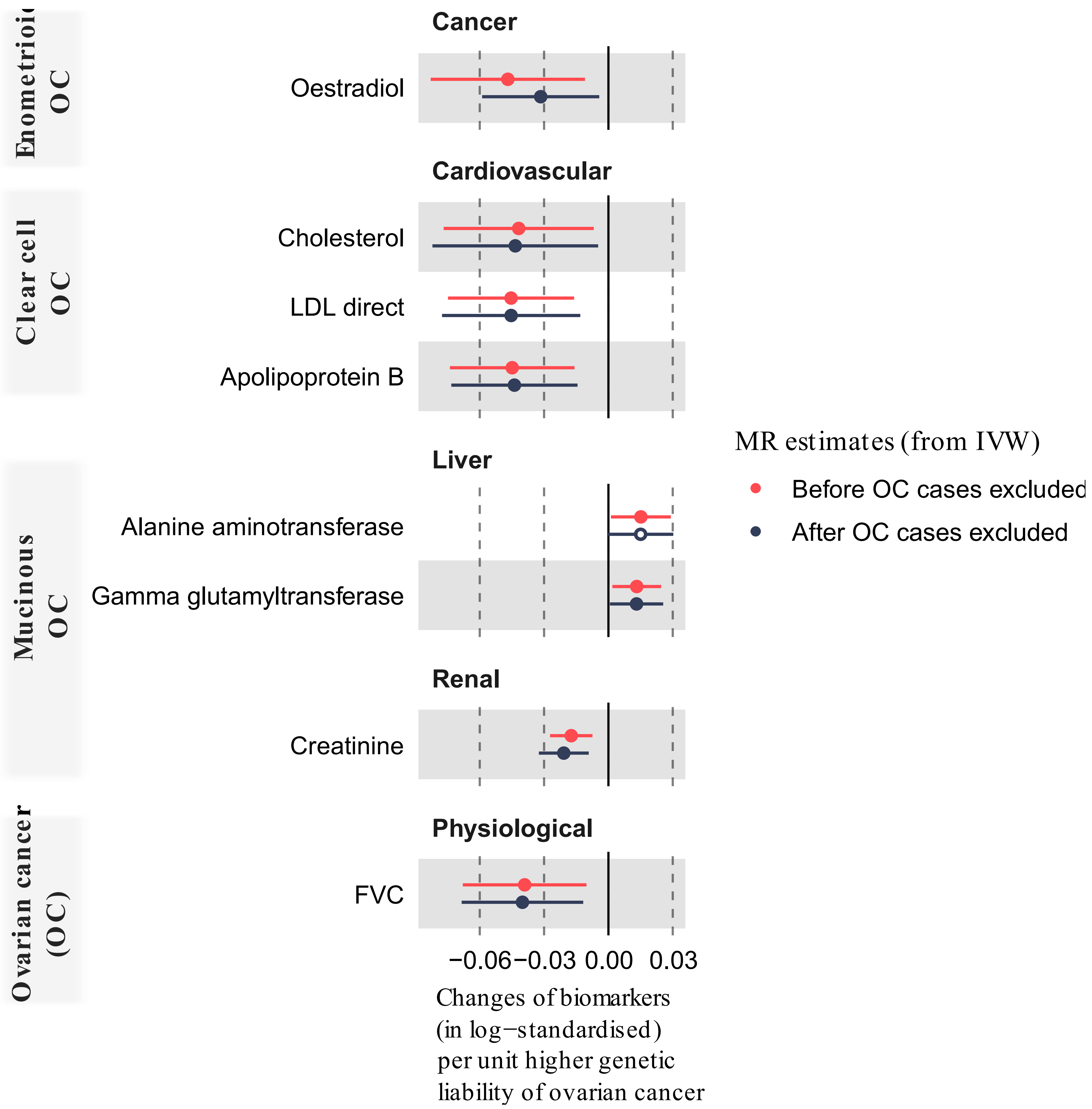


**Figure S11.** Mendelian randomisation estimates before (main analysis) and after removing (sensitivity analysis) ovarian cancer cases.

All biomarkers were log-transformed and standardised to mean zero and standard deviation of one.

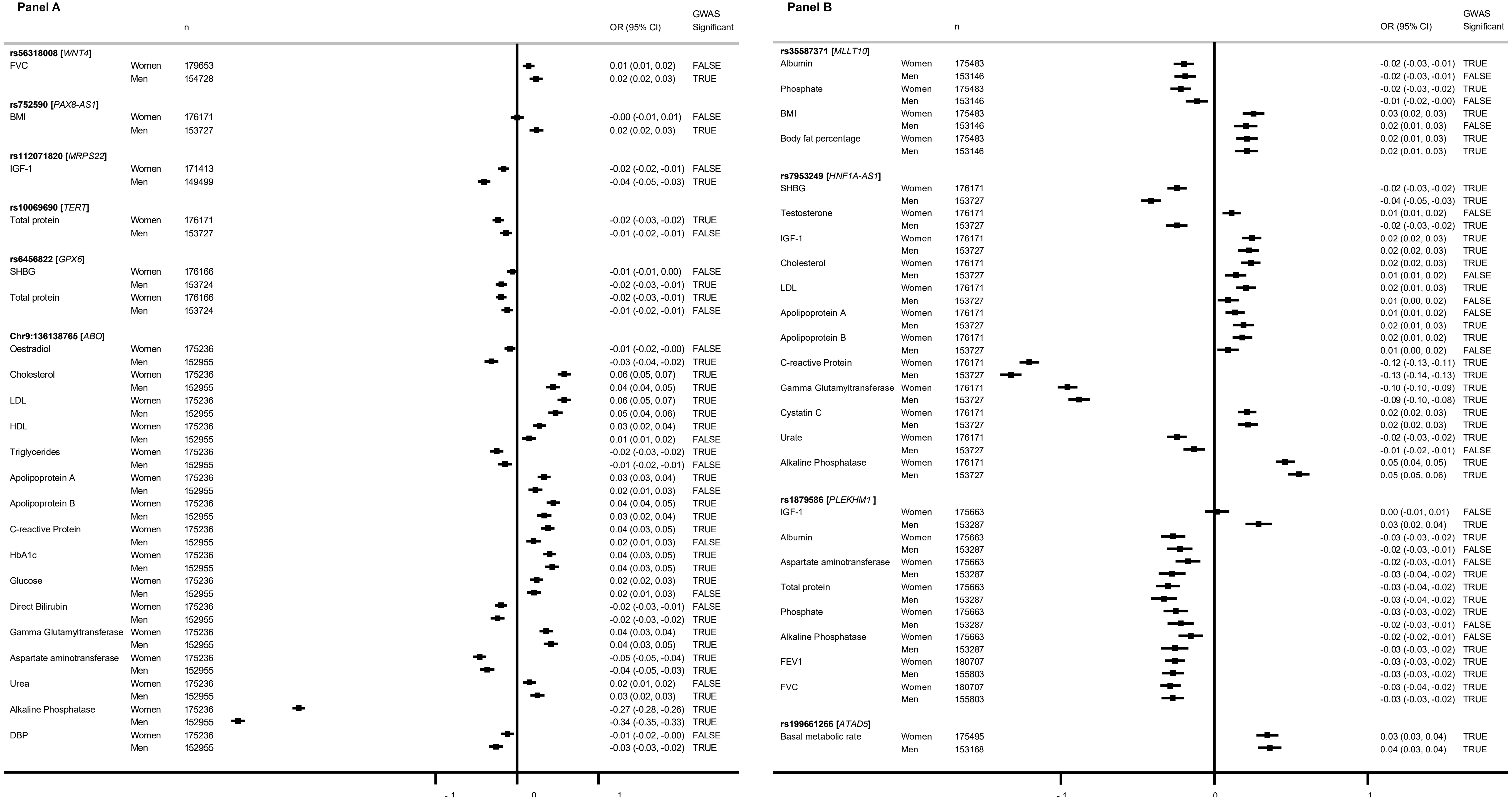

**Panel A**

| | | n | OR (95% CI) | GWAS Significant |
|---|---|---|---|---|
| **rs56318008 [*WNT4*]** | | | | |
| FVC | Women | 179653 | 0.01 (0.01, 0.02) | FALSE |
| | Men | 154728 | 0.02 (0.02, 0.03) | TRUE |
| **rs752590 [*PAX8-AS1*]** | | | | |
| BMI | Women | 176171 | -0.00 (-0.01, 0.01) | FALSE |
| | Men | 153727 | 0.02 (0.02, 0.03) | TRUE |
| **rs112071820 [*MRPS22*]** | | | | |
| IGF-1 | Women | 171413 | -0.02 (-0.02, -0.01) | FALSE |
| | Men | 149499 | -0.04 (-0.05, -0.03) | TRUE |
| **rs10069690 [*TERT*]** | | | | |
| Total protein | Women | 176171 | -0.02 (-0.03, -0.02) | TRUE |
| | Men | 153727 | -0.01 (-0.02, -0.01) | FALSE |
| **rs6456822 [*GPX6*]** | | | | |
| SHBG | Women | 176166 | -0.01 (-0.01, 0.00) | FALSE |
| | Men | 153724 | -0.02 (-0.03, -0.01) | TRUE |
| Total protein | Women | 176166 | -0.02 (-0.03, -0.01) | TRUE |
| | Men | 153724 | -0.01 (-0.02, -0.01) | FALSE |
| **Chr9:136138765 [*ABO*]** | | | | |
| Oestradiol | Women | 175236 | -0.01 (-0.02, -0.00) | FALSE |
| | Men | 152955 | -0.03 (-0.04, -0.02) | TRUE |
| Cholesterol | Women | 175236 | 0.06 (0.05, 0.07) | TRUE |
| | Men | 152955 | 0.04 (0.04, 0.05) | TRUE |
| LDL | Women | 175236 | 0.06 (0.05, 0.07) | TRUE |
| | Men | 152955 | 0.05 (0.04, 0.06) | TRUE |
| HDL | Women | 175236 | 0.03 (0.02, 0.04) | TRUE |
| | Men | 152955 | 0.01 (0.01, 0.02) | FALSE |
| Triglycerides | Women | 175236 | -0.02 (-0.03, -0.02) | TRUE |
| | Men | 152955 | -0.01 (-0.02, -0.01) | FALSE |
| Apolipoprotein A | Women | 175236 | 0.03 (0.03, 0.04) | TRUE |
| | Men | 152955 | 0.02 (0.01, 0.03) | FALSE |
| Apolipoprotein B | Women | 175236 | 0.04 (0.04, 0.05) | TRUE |
| | Men | 152955 | 0.03 (0.02, 0.04) | TRUE |
| C-reactive Protein | Women | 175236 | 0.04 (0.03, 0.05) | TRUE |
| | Men | 152955 | 0.02 (0.01, 0.03) | FALSE |
| HbA1c | Women | 175236 | 0.04 (0.03, 0.05) | TRUE |
| | Men | 152955 | 0.04 (0.03, 0.05) | TRUE |
| Glucose | Women | 175236 | 0.02 (0.02, 0.03) | TRUE |
| | Men | 152955 | 0.02 (0.01, 0.03) | FALSE |
| Direct Bilirubin | Women | 175236 | -0.02 (-0.03, -0.01) | FALSE |
| | Men | 152955 | -0.02 (-0.03, -0.02) | TRUE |
| Gamma Glutamyltransferase | Women | 175236 | 0.04 (0.03, 0.04) | TRUE |
| | Men | 152955 | 0.04 (0.03, 0.05) | TRUE |
| Aspartate aminotransferase | Women | 175236 | -0.05 (-0.05, -0.04) | TRUE |
| | Men | 152955 | -0.04 (-0.05, -0.03) | TRUE |
| Urea | Women | 175236 | 0.02 (0.01, 0.02) | FALSE |
| | Men | 152955 | 0.03 (0.02, 0.03) | TRUE |
| Alkaline Phosphatase | Women | 175236 | -0.27 (-0.28, -0.26) | TRUE |
| | Men | 152955 | -0.34 (-0.35, -0.33) | TRUE |
| DBP | Women | 175236 | -0.01 (-0.02, -0.00) | FALSE |
| | Men | 152955 | -0.03 (-0.03, -0.02) | TRUE |

**Panel B**

| | | n | OR (95% CI) | GWAS Significant |
|---|---|---|---|---|
| **rs35587371 [*MLLT10*]** | | | | |
| Albumin | Women | 175483 | -0.02 (-0.03, -0.01) | TRUE |
| | Men | 153146 | -0.02 (-0.03, -0.01) | FALSE |
| Phosphate | Women | 175483 | -0.02 (-0.03, -0.02) | TRUE |
| | Men | 153146 | -0.01 (-0.02, -0.00) | FALSE |
| BMI | Women | 175483 | 0.03 (0.02, 0.03) | TRUE |
| | Men | 153146 | 0.02 (0.01, 0.03) | FALSE |
| Body fat percentage | Women | 175483 | 0.02 (0.01, 0.03) | TRUE |
| | Men | 153146 | 0.02 (0.01, 0.03) | TRUE |
| **rs7953249 [*HNF1A-AS1*]** | | | | |
| SHBG | Women | 176171 | -0.02 (-0.03, -0.02) | TRUE |
| | Men | 153727 | -0.04 (-0.05, -0.03) | TRUE |
| Testosterone | Women | 176171 | 0.01 (0.01, 0.02) | FALSE |
| | Men | 153727 | -0.02 (-0.03, -0.02) | TRUE |
| IGF-1 | Women | 176171 | 0.02 (0.02, 0.03) | TRUE |
| | Men | 153727 | 0.02 (0.02, 0.03) | TRUE |
| Cholesterol | Women | 176171 | 0.02 (0.02, 0.03) | TRUE |
| | Men | 153727 | 0.01 (0.01, 0.02) | FALSE |
| LDL | Women | 176171 | 0.02 (0.01, 0.03) | TRUE |
| | Men | 153727 | 0.01 (0.00, 0.02) | FALSE |
| Apolipoprotein A | Women | 176171 | 0.01 (0.01, 0.02) | FALSE |
| | Men | 153727 | 0.02 (0.01, 0.03) | TRUE |
| Apolipoprotein B | Women | 176171 | 0.02 (0.01, 0.02) | TRUE |
| | Men | 153727 | 0.01 (0.00, 0.02) | FALSE |
| C-reactive Protein | Women | 176171 | -0.12 (-0.13, -0.11) | TRUE |
| | Men | 153727 | -0.13 (-0.14, -0.13) | TRUE |
| Gamma Glutamyltransferase | Women | 176171 | -0.10 (-0.10, -0.09) | TRUE |
| | Men | 153727 | -0.09 (-0.10, -0.08) | TRUE |
| Cystatin C | Women | 176171 | 0.02 (0.02, 0.03) | TRUE |
| | Men | 153727 | 0.02 (0.02, 0.03) | TRUE |
| Urate | Women | 176171 | -0.02 (-0.03, -0.02) | TRUE |
| | Men | 153727 | -0.01 (-0.02, -0.01) | FALSE |
| Alkaline Phosphatase | Women | 176171 | 0.05 (0.04, 0.05) | TRUE |
| | Men | 153727 | 0.05 (0.05, 0.06) | TRUE |
| **rs1879586 [*PLEKHM1* ]** | | | | |
| IGF-1 | Women | 175663 | 0.00 (-0.01, 0.01) | FALSE |
| | Men | 153287 | 0.03 (0.02, 0.04) | TRUE |
| Albumin | Women | 175663 | -0.03 (-0.03, -0.02) | TRUE |
| | Men | 153287 | -0.02 (-0.03, -0.01) | FALSE |
| Aspartate aminotransferase | Women | 175663 | -0.02 (-0.03, -0.01) | FALSE |
| | Men | 153287 | -0.03 (-0.04, -0.02) | TRUE |
| Total protein | Women | 175663 | -0.03 (-0.04, -0.02) | TRUE |
| | Men | 153287 | -0.03 (-0.04, -0.02) | TRUE |
| Phosphate | Women | 175663 | -0.03 (-0.03, -0.02) | TRUE |
| | Men | 153287 | -0.02 (-0.03, -0.01) | FALSE |
| Alkaline Phosphatase | Women | 175663 | -0.02 (-0.02, -0.01) | FALSE |
| | Men | 153287 | -0.03 (-0.03, -0.02) | TRUE |
| FEV1 | Women | 180707 | -0.03 (-0.03, -0.02) | TRUE |
| | Men | 155803 | -0.03 (-0.03, -0.02) | TRUE |
| FVC | Women | 180707 | -0.03 (-0.04, -0.02) | TRUE |
| | Men | 155803 | -0.03 (-0.03, -0.02) | TRUE |
| **rs199661266 [*ATAD5*]** | | | | |
| Basal metabolic rate | Women | 175495 | 0.03 (0.03, 0.04) | TRUE |
| | Men | 153168 | 0.04 (0.03, 0.04) | TRUE |

**Figure S12**. Phenome-wide associations between OC associated variants and biomarkers and physiological measures

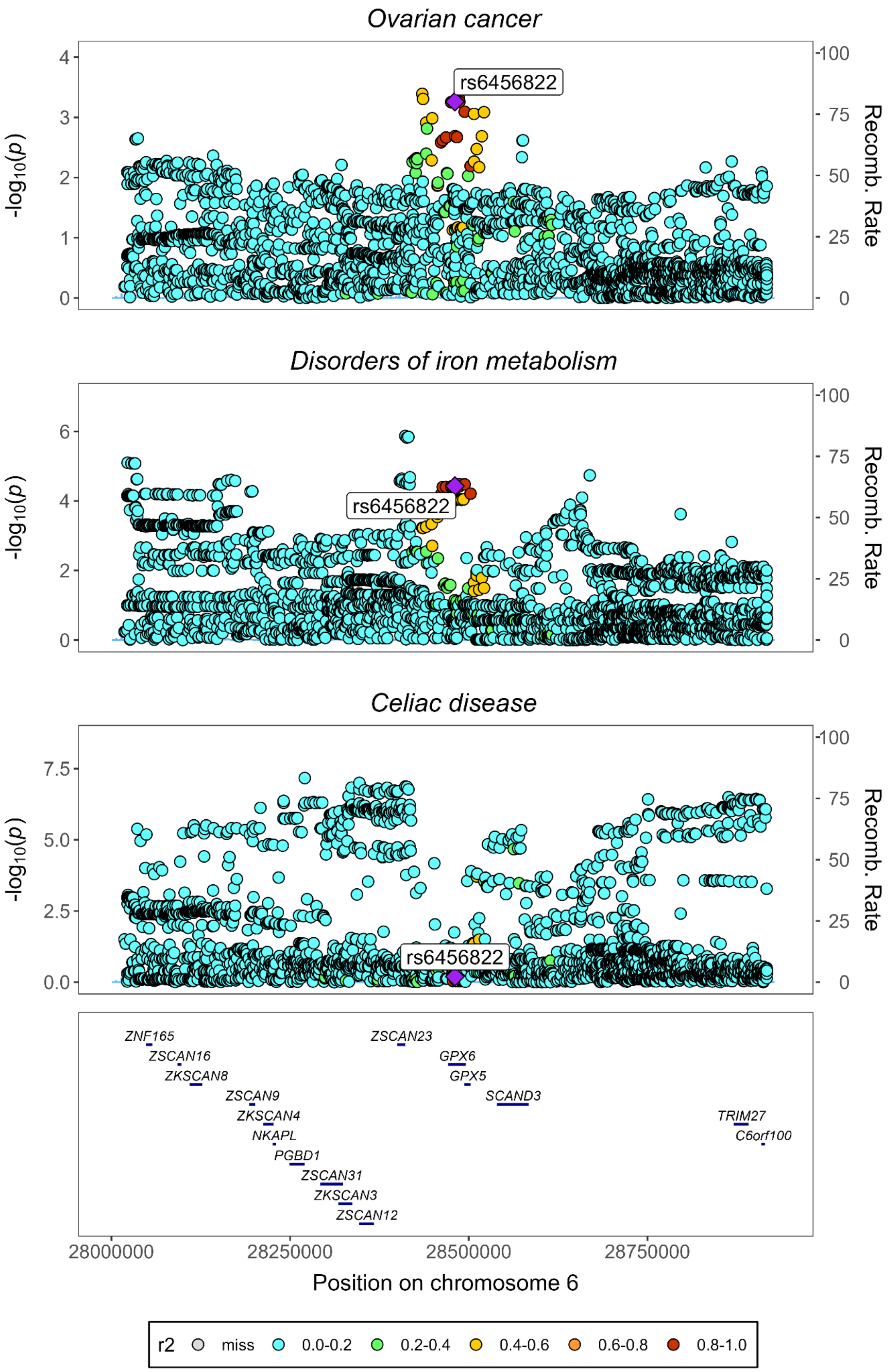


**Figure S13**. Regional association plots for ovarian cancer, disorders of iron metabolism and celiac disease at *GPX6* locus (Chr6: 28017819 – Chr6:28917608, hg19), after conditional analysis for the lead variant of ovarian cancer (rs6456822), disorder of iron metabolism (rs34409925) and celiac disease (rs3131101).
